# Derivation and external validation of a simple risk score to predict in-hospital mortality in patients hospitalized for COVID-19

**DOI:** 10.1101/2021.05.04.21256599

**Authors:** Charlotte Z. Mann, Chelsea Abshire, Monica Yost, Scott Kaatz, Lakshmi Swaminathan, Scott A. Flanders, Hallie C. Prescott, Johann A. Gagnon-Bartsch

## Abstract

**Background:** As SARS-CoV-2 continues to spread, and hospitals are treating a large number of patients with COVID-19, easy-to-use risk models that predict hospital mortality can assist in clinical decision making and triage.

**Objective:** As SARS-CoV-2 continues to spread, easy-to-use risk models that predict hospital mortality can assist in clinical decision making and triage. We aimed to develop a risk score model for in-hospital mortality in patients hospitalized with COVID-19 that was robust across hospitals and used clinical factors that are readily available and measured standardly across hospitals.

**Methods:** In this observational study we developed a risk score model using data collected by trained abstractors for patients in 20 diverse hospitals across the state of Michigan (Mi-COVID19) who were discharged between March 5, 2020 and August 14, 2020. Patients who tested positive for SARS-CoV-2 during hospitalization or were discharged with an ICD-10 code for COVID-19 (U07.1) were included. We employed an iterative forward selection approach to consider the inclusion of 145 potential risk factors available at hospital presentation. Model performance was externally validated with patients from 19 hospitals in the Mi-COVID19 registry not used in model development. We shared the model in an easy-to-use online application that allows the user to predict in-hospital mortality risk for a patient if they have any subset of the variables in the final model.

**Results:** Our final model includes the patient’s age, first recorded respiratory rate, first recorded pulse oximetry, highest creatinine level on day of presentation, and hospital’s COVID-19 mortality rate. No other factors showed sufficient incremental model improvement to warrant inclusion. The AUC for the derivation and validation sets were .796 and .829 respectively.

**Conclusions:** Risk of in-hospital mortality in COVID-19 patients can be reliably estimated using a few factors, which are standardly measured and available to physicians very early in a hospital encounter.

## 1. Introduction

The COVID-19 outbreak was declared a pandemic by the World Health Organization in March 2020 and continues to devastate much of the world. As of April 18, 2021, there have been approximately 141 million reported COVID-19 cases worldwide and 3.02 million reported deaths attributed to COVID-19, with a mortality rate per reported infection of 2.1%^1^. During the pandemic, hospitals have needed to constantly adapt how they operate as COVID-19 cases rise and wane. Empirical tools to assess individual patients’ risk for mortality could be lifesaving in hospitals, where decisions must be made as to how to allocate resources. To fill this need, since March 2020, a number of risk score models for predicting adverse outcomes of COVID-19 have been published^2–5^. However, many of these models were developed in patients within a single hospital or hospital system, lacked validation, or incorporated variables that are not routinely available or not measured consistently across hospitals or required subjective evaluation. Concerns of generalizability and ease of implementation remain.

In this study, we aimed to develop a model to predict the risk of in-hospital mortality among patients hospitalized for COVID-19, utilizing variables that are readily available when a patient is hospitalized. We used a systematic variable selection approach to determine a small number of highly predictive factors from over one hundred variables in a high-quality observational dataset abstracted from patient electronic health records in the state of Michigan.

## 2. Methods

### 2.1 Study Design and Data Abstraction

We used data abstracted from patients in hospitals across the state of Michigan (Mi-COVID19 registry) to fit a model predictive of risk of in-hospital death amongst patients hospitalized for COVID-19.^6^ The Mi-COVID19 data registry is a joint initiative between 10 collaborative quality initiatives sponsored by an insurance provider, Blue Cross Blue Shield of Michigan and Blue Care Network to create a multi-hospital data registry.^6^ Forty non-critical access, non-federal Michigan hospitals voluntarily participated in abstracting data from patients, following a coordinated abstraction protocol, beginning on April 2, 2020.

Data were abstracted from medical records by trained abstractors. The data included a pseudo-random sample of COVID-19 positive cases discharged between March 5, 2020 and August 14, 2020 (with the majority of discharges before April 24, 2020). If a hospital had the ability to abstract full patient data for all eligible COVID-19 positive patients that entered the hospital, they did so. However, if a hospital was unable to abstract all cases, each day the eligible cases were ordered by the timestamp (minute) of discharge and were included starting with the smallest minute value of discharge, until the hospital reached their abstraction capacity.

For this study, we only included patients who tested positive for SARS-CoV-2 within the hospital in which they were enrolled or were discharged with an ICD-10 code for COVID-19 (U07.1). We split the data to include 20 hospitals in a derivation set and 20 hospitals in a validation set. Our derivation set included the 20 hospitals with the largest number of patients with abstracted data and we reserved the data on patients in the 20 hospitals with the smallest sample sizes for the validation set. We split the data by hospitals, rather than taking a random sample of patients, to emulate external validation and ensure that our estimates of model discrimination were not overly optimistic. We used a complete case analysis, so we ultimately excluded one hospital in the validation set since it had no observations with complete cases for the variables included in our final model. It is worth noting that the sample size available in the Mi-COVID19 data registry did not necessarily correlate with the size of the hospital, so there are small and large hospitals in both the derivation and validation sets.

### 2.2 Outcome and Potential Risk Factors

The outcome of interest was in-hospital mortality. We considered as possible risk factors 145 variables abstracted in Mi-COVID19 which were measured on the first or second days of hospitalization. These risk factors included a patient’s 1) demographics and health behaviors (e.g., smoking), 2) medical history (comorbidities and previous medications and treatments), 3) exposure risk factors (e.g., being a service or healthcare worker), 4) symptoms and primary complaint at hospital arrival, 5) triage and first day vital signs, 6) first or second hospitalization day lab values 7) first or second hospitalization day chest x-ray findings. See eTable 1, Supplemental Content, for a full list of factors considered in model derivation.

### 2.3 Statistical Methods

We predicted in-hospital mortality using a logistic regression model. Our approach to variable selection ensured that our model was robust across the hospitals participating in Mi-COVID19 and used factors that are commonly measured, while maintaining high discrimination. In order to control for variability between hospitals, we set our base model to be each hospital’s COVID-19 mortality rate, which can be thought of as the patient’s pre-test probability for mortality at a given hospital. We included the hospital’s COVID-19 mortality rate as an adjustment to ensure that hospital-level differences in mortality would not be falsely attributed to patient characteristics or mask important patient characteristics. From this base model, we used forward and backward selection to choose which variables to include in the model.

In the forward and backward selection, we assessed the model with three quality metrics: mean squared error (MSE), R-squared, and an adjusted area under the receiver operating characteristic curve (AUC). The adjusted AUC (denoted AUC^(w)^) assessed the discrimination of the model in a way that was unaffected by the value of the mortality rate of each hospital (see eMethods, Supplemental Content, for details on this calculation). We used this metric so that the AUC was not falsely inflated by including hospital mortality rate in the model. The value of AUC^(w)^ can be interpreted as an estimate of the probability that, for two randomly selected patients with opposite outcomes from the same hospital, the model estimates a higher risk of mortality for the patient who is deceased. The AUC^(w)^ calculation is a weighted average of the individual hospital AUCs, with weights proportional to the sample size. If AUC^(w)^ is calculated for only one hospital, it is equivalent to the standard AUC.

For each step of forward selection, we added every potential risk factor individually to the model determined from the previous step. We then assessed how much the model’s mean squared error (MSE), AUC^(w)^, and R-squared improved when including a variable as compared to the model excluding that variable. We assessed improvement in the quality metrics (MSE, AUC^(w)^, and R-squared) both for the full derivation dataset (all 20 hospitals) and within each hospital individually. We considered adding variables to the base model only if there was consistent improvement in MSE, AUC^(w)^, and R-squared across the individual hospitals as well as all hospitals combined and if the improvement across all derivation hospitals was sufficiently large to warrant inclusion. Amongst factors that had consistent quantitative support to be added to the model, we considered whether these factors were likely to be routinely available and standardly reported across hospitals, as inferred by our clinical experience. If the answer was “yes,” we added the variable to the model. We repeated this process until there were no remaining clinically meaningful and standardly measured variables that also dependably improved MSE, AUC^(w)^, and R-squared in a meaningful way across hospitals in the derivation set when added to the model.

Following this forward selection protocol, we additionally performed one step of backward selection to ensure that all variables, given all others in the model, were predictive of in-hospital mortality. We removed each variable individually and again assessed the change in MSE, AUC^(w)^, and R-squared from the initial model to this model with one variable removed. If the MSE consistently increased and the AUC^(w)^ and R-squared consistently decreased across hospitals when the variable was removed from the model, then we kept that variable in our final model.

In both forward and backward selection, predictions for each individual hospital were made using a model fit on all other hospitals in the derivation set. In this way, all models were assessed using leave-one-out predictions, which reduced the concern for overfitting.

### 2.4 Model Validation

We assessed the performance of our final model on the external validation set of 19 hospitals described previously. We additionally assessed the performance of the model across subgroups of patients (Black versus white, male versus female, and 75 years of age or older versus younger than 75) in terms of AUC for the full dataset.

### 2.5 Model Application

We aimed to develop a mortality risk assessment model that was as accessible as possible; therefore, we shared the model using an online app developed in R (see eFigure 1, Supplemental Content). One can input the values for the patient characteristics included in the final model into the app and the predicted risk of mortality is outputted. We set up the app such that one does not need to have or guess the values of every variable used in our final risk score model in order to estimate a patient’s risk of mortality. Instead, the app will refit the model using variables that the user does have access to (see eMethods, Supplemental Content, for details). The app can be accessed at https://micovidriskcalc.org/.

The study was deemed exempt by the University of Michigan Institutional Review Board. Data management and analysis was completed in SAS and R version 4.0.3.

## 3. Results

### 3.1 Patient and Hospital Characteristics

The Mi-Covid19 dataset included 2,193 patients who met our inclusion criteria. In the final model validation, we excluded 79 (4.5%) individuals in the derivation set and 26 (6.1%) individuals in the validation set who were missing data for one of the predictive factors in the final model. Therefore, our final derivation and validation sets included 1,690 and 398 patients, amongst the 20 and 19 hospitals respectively. The demographic and clinical characteristics of the patients in the derivation set are described in Table 1. See eTable 2 and eTable 3, Supplemental Content, for hospital characteristics in the Mi-COVID19 registry and characteristics of the patients in the validation set. The overall in-hospital mortality rates in the derivation and validations sets were 19.6% and 18.6% respectively. However, there was variability of mortality rates between individual hospitals. The mortality rates in individual derivation set hospitals ranged from 7.4% to 54.3% (see eFigure 2 and eTable 2, Supplemental Content, for all hospital COVID-19 mortality rates).

**Table 1:** Derivation set patient characteristics.

| Characteristic | Overall<br>[N = 1769] | In-Hospital Mortality |  |
| --- | --- | --- | --- |
|  | mean/No. (SD/%)<br>[n*] | No<br>[N = 1422] | Yes<br>[N = 347] |
| Age | 64.4 (16.7) [1769] | 62 (16.6) [1422] | 74.4 (13.1) [347] |
| Gender (female) | 830 (47%) [1769] | 670 (47%) [1422] | 160 (46%) [347] |
| Race (yes) |  |  |  |
| Black | 840 (49%) [1698] | 687 (50%) [1364] | 153 (46%) [334] |
| White | 744 (44%) [1698] | 579 (42%) [1364] | 165 (49%) [334] |
| Asian | 45 (3%) [1698] | 38 (3%) [1364] | 7 (2%) [334] |
| Native American or Pacific Islander | 10 (1%) [1698] | 8 (1%) [1364] | 2 (1%) [334] |
| Other | 59 (3%) [1698] | 52 (4%) [1364] | 7 (2%) [334] |
| Ethnicity (yes) |  |  |  |
| Hispanic | 92 (5%) [1762] | 84 (6%) [1415] | 8 (2%) [347] |
| Non-Hispanic | 1535 (87%) [1762] | 1229 (87%) [1415] | 306 (88%) [347] |
| Unknown | 135 (8%) [1762] | 102 (7%) [1415] | 33 (10%) [347] |
| Residing in a Nursing Facility or Assisted Living (yes) | 340 (19%) [1748] | 207 (15%) [1406] | 133 (39%) [342] |
| Ever-smoker (yes) | 645 (39%) [1648] | 515 (38%) [1355] | 130 (44%) [293] |
| BMI | 31.2 (8.5) [1680] | 31.5 (8.4) [1359] | 30 (8.8) [321] |
| No. of comorbidities |  |  |  |
| 0 | 228 (13%) [1769] | 211 (15%) [1422] | 17 (5%) [347] |
| 1 | 341 (19%) [1769] | 314 (22%) [1422] | 27 (8%) [347] |
| 2 | 369 (21%) [1769] | 309 (22%) [1422] | 60 (17%) [347] |
| 3 | 299 (17%) [1769] | 223 (16%) [1422] | 76 (22%) [347] |
| 4 | 225 (13%) [1769] | 166 (12%) [1422] | 59 (17%) [347] |
| >4 | 307 (17%) [1769] | 199 (14%) [1422] | 108 (31%) [347] |
| Presence of comorbidity (yes) |  |  |  |
| Cardiovascular disease | 485 (27%) [1769] | 347 (24%) [1422] | 138 (40%) [347] |
| Congestive heart failure | 275 (16%) [1769] | 191 (13%) [1422] | 84 (24%) [347] |
| Chronic obstructive pulmonary disease | 208 (12%) [1769] | 149 (10%) [1422] | 59 (17%) [347] |
| Asthma | 219 (12%) [1769] | 189 (13%) [1422] | 30 (9%) [347] |
| Diabetes (complicated and uncomplicated) | 655 (37%) [1769] | 487 (34%) [1422] | 168 (48%) [347] |
| Severe liver disease | 12 (1%) [1769] | 10 (1%) [1422] | 2 (1%) [347] |
| Cancer | 144 (8%) [1769] | 106 (7%) [1422] | 38 (11%) [347] |
| Symptoms (yes) |  |  |  |
| Fatigue | 585 (33%) [1769] | 491 (35%) [1422] | 94 (27%) [347] |
| Fever (subjective and objective) | 1452 (82%) [1769] | 1204 (85%) [1422] | 248 (71%) [347] |
| Chest pain | 301 (17%) [1769] | 263 (18%) [1422] | 38 (11%) [347] |
| Hypoxia | 729 (41%) [1769] | 526 (37%) [1422] | 203 (59%) [347] |
| First recorded heart rate |  |  |  |
| < 90 BPM | 701 (40%) [1760] | 572 (40%) [1415] | 129 (37%) [345] |
| 90-100 BPM | 391 (22%) [1760] | 321 (23%) [1415] | 70 (20%) [345] |
| 101-124 BPM | 544 (31%) [1760] | 442 (31%) [1415] | 102 (30%) [345] |
| > 124 BPM | 124 (7%) [1760] | 80 (6%) [1415] | 44 (13%) [345] |
| First recorded respiratory rate |  |  |  |
| < 20 | 645 (37%) [1734] | 564 (41%) [1390] | 81 (24%) [344] |
| 20-24 | 682 (39%) [1734] | 559 (40%) [1390] | 123 (36%) [344] |
| 25-30 | 240 (14%) [1734] | 173 (12%) [1390] | 67 (19%) [344] |
| > 30 | 167 (10%) [1734] | 94 (7%) [1390] | 73 (21%) [344] |

| Characteristic | Overall<br>[N = 1769]<br>mean/No. (SD/%)<br>[n*] | In-Hospital Mortality |  |
| --- | --- | --- | --- |
|  |  | No<br>[N = 1422] | Yes<br>[N = 347] |
| First recorded systolic blood pressure |  |  |  |
| >= 101 mmHg | 1619 (93%) [1740] | 1318 (94%) [1401] | 301 (89%) [339] |
| 90 - 100 mmHg | 80 (5%) [1740] | 52 (4%) [1401] | 28 (8%) [339] |
| < 90 mmHg | 41 (2%) [1740] | 31 (2%) [1401] | 10 (3%) [339] |
| First recorded pulse oximetry |  |  |  |
| 91-100% | 1386 (79%) [1750] | 1147 (82%) [1406] | 239 (69%) [344] |
| 81-90% | 287 (16%) [1750] | 219 (16%) [1406] | 68 (20%) [344] |
| 71-80% | 45 (3%) [1750] | 26 (2%) [1406] | 19 (6%) [344] |
| <= 70% | 32 (2%) [1750] | 14 (1%) [1406] | 18 (5%) [344] |
| Triage score |  |  |  |
| 1 | 91 (6%) [1564] | 43 (3%) [1259] | 48 (16%) [305] |
| 2 | 731 (47%) [1564] | 570 (45%) [1259] | 161 (53%) [305] |
| 3 | 646 (41%) [1564] | 582 (46%) [1259] | 64 (21%) [305] |
| 4 | 37 (2%) [1564] | 35 (3%) [1259] | 2 (1%) [305] |
| 5 | 59 (4%) [1564] | 29 (2%) [1259] | 30 (10%) [305] |
| Highest initial creatinine (mg/dL) | 1.7 (1.7) [1737] | 1.5 (1.6) [1392] | 2.3 (2) [345] |
| Highest initial white blood cell count (K/uL) | 8.4 (6.7) [1750] | 8.1 (6.9) [1405] | 9.9 (5.5) [345] |
| Pneumonia indication on chest x-ray (yes) | 1318 (78%) [1684] | 1024 (77%) [1337] | 294 (85%) [347] |

\* n is the number of complete cases in the data for the given variable. Percentages are calculated as No./n. SD = Standard Deviation, BPM = Beats per Minute, BMI = Body Mass Index.

### 3.2 Risk Score Model

Through our forward and backward variable selection process, we arrived at a final risk score model that included patient age, first recorded pulse oximetry, first recorded respiratory rate, and highest creatinine level on the first day of admission, along with the overall mortality rate at the patient’s hospital. The first variable that met the forward selection criteria was patient age, then pulse oximetry, respiratory rate, and heart rate in the second step of forward selection, and then finally creatinine in the third step. Then, based on the backward selection criteria, we removed heart rate. In a final step of forward selection, no further factors met our criteria. See eAppendix A and eFigure 3, Supplemental Content, for details.

During the forward selection process, the manner in which a patient arrived at the ED (e.g., by ambulance or by foot) seemed to improve the model (improving AUC^(w)^ by 2-13% in the first steps of forward selection), however this data was not reliably available in different hospitals. The interleukin-6 and creatine phosphokinase labs additionally appeared to improve the model in terms of MSE (improving MSE by .003, the most out of other variables in the second and third steps of forward selection), however these lab values were available for only 156 (7%) and 562 (26%) patients of the patients in the MiCOVID-19 data, respectively, and are unlikely to be commonly measured. Finally, the patient’s emergency department triage score (a number from 1-5 indicating acuity with 1 as highest acuity and 5 as lowest acuity) and the presence of hypoxia as a symptom appeared predictive in the first steps of forward selection (improving the AUC^(w)^ by 3%), however we chose to add vital signs before symptoms since they have objectively measured values and before the triage score because the vital signs are incorporated in that score. Once the vital signs were added to the model, the marginal gain from adding hypoxia and triage score (only improving AUC^(w)^ by .7-1%) was not sufficient to warrant a more complicated model (see eAppendix A, Supplemental Content). A number of factors that have appeared in other risk models did not appear predictive in our model after controlling for the hospital, for instance a patient’s BMI, race, and gender.

Thus, we predict risk of in-hospital mortality using a logistic regression model with five covariates. We applied a spline basis transformation to age, with knots at 35, 50, 65, and 80 years, to allow for a non-linear relationship with risk, and a log transformation to creatinine level. Pulse oximetry on admission was collected as “70% or less,” “71-80%,” “81-90%,” and “91-100%.” Respiratory rate was collected as “Abnormal (20-24),” “Abnormal (25 - 30),” “Abnormal (greater than 30),” “Normal (less than 20).” Table 2 reports the odds ratios corresponding to the coefficients of this model, fit on the derivation set (see eFigure 4, Supplemental Content, for the age odds ratios). In a logistic regression model, the odds (i.e., the probability of mortality divided by the probability of survival) is estimated by *e*^*β*^′^*X*^ where β is a vector of coefficients and X is the matrix of covariates. We estimate that the odds of mortality increases as age, respiratory rate, and creatinine increase, and as pulse oximetry decreases. See Table 3 for examples of the predicted risk of mortality for twelve patients with different characteristics.

**Table 2:** Odds ratios associated with coefficients of final risk score model.

| Factor | Odds Ratio | 95% CI | p |
| --- | --- | --- | --- |
| **Age\*** |  |  |  |
| 50 (referent) | 1.00 |  |  |
| 30 | 0.28 | (0.09-0.87) |  |
| 40 | 0.53 | (0.32-0.87) |  |
| 60 | 1.87 | (1.58-2.22) |  |
| 70 | 3.26 | (2.64-4.02) |  |
| 80 | 5.26 | (4.07-6.81) |  |
| **Respiratory Rate** |  |  |  |
| less than 20 (referent) | 1.00 |  |  |
| 20-24 | 1.50 | (1.06-2.12) | 0.022 |
| 25-30 | 2.57 | (1.68-3.93) | <.001 |
| greater than 30 | 3.88 | (2.42-6.23) | <.001 |
| **Pulse Oximetry** |  |  |  |
| 91-100% (referent) | 1.00 |  |  |
| 81-90% | 1.44 | (1.00-2.07) | 0.05 |
| 71-80% | 3.66 | (1.81-7.42) | <.001 |
| 70% or less | 5.57 | (2.22-13.92) | <.001 |
| **Creatinine (log transformed)** | 2.29 | (1.85-2.84) | <.001 |
| **Hospital mortality rate (logit transformed)** | 2.81 | (2.28-3.45) | <.001 |
| **Constant** | 0.06 | (0.02-0.14) | <.001 |

\* We applied a spline transformation on age, however the spline regression coefficients are not interpretable. Therefore, in this table we display the estimated odds ratios for different ages as compared to 50 years as a reference age. The coefficients for the spline are significant with p-values <.001. See eFigure 4 for an illustration of the odds ratios and confidence intervals for age with 50 years as a reference age for the full range of ages in the data.

**Table 3:** Estimated risk of in-hospital mortality for ten example patients.

| Example Patient | Creatinine (mg/dL) | Age (years) | Respiratory Rate | Pulse Oximetry | Estimated Risk |
| --- | --- | --- | --- | --- | --- |
| Patient 1 | 1 | 50 | 20 | 82% | 7% |
| Patient 2 | 1 | 50 | 20 | 92% | 5% |
| Patient 3 | 1 | 50 | 30 | 82% | 11% |
| Patient 4 | 1 | 50 | 30 | 92% | 8% |
| Patient 5 | 1 | 75 | 20 | 82% | 24% |
| Patient 6 | 1 | 75 | 20 | 92% | 18% |
| Patient 7 | 1 | 75 | 30 | 82% | 35% |
| Patient 8 | 1 | 75 | 30 | 92% | 27% |
| Patient 9 | 2 | 50 | 20 | 92% | 8% |
| Patient 10 | 2 | 75 | 20 | 92% | 28% |

### 3.3 Model Validation

The AUC^(w)^ for the model on the derivation set was .796. The model had similar discrimination on the validation set, with an AUC^(w)^ of .829. The individual hospital AUC^(w)^ values for the validation set vary around .83 and show good discrimination (see Figure 1). We also found that the model shows similar discrimination for Black and white patients as well as male and female patients, although the discrimination is not as strong for patients 75 years or older (see eAppendix B and eTable 4, Supplemental Content). Finally, we observe good model calibration when using the individual hospital mortality rates (see Figure 2).

**Figure 1:**
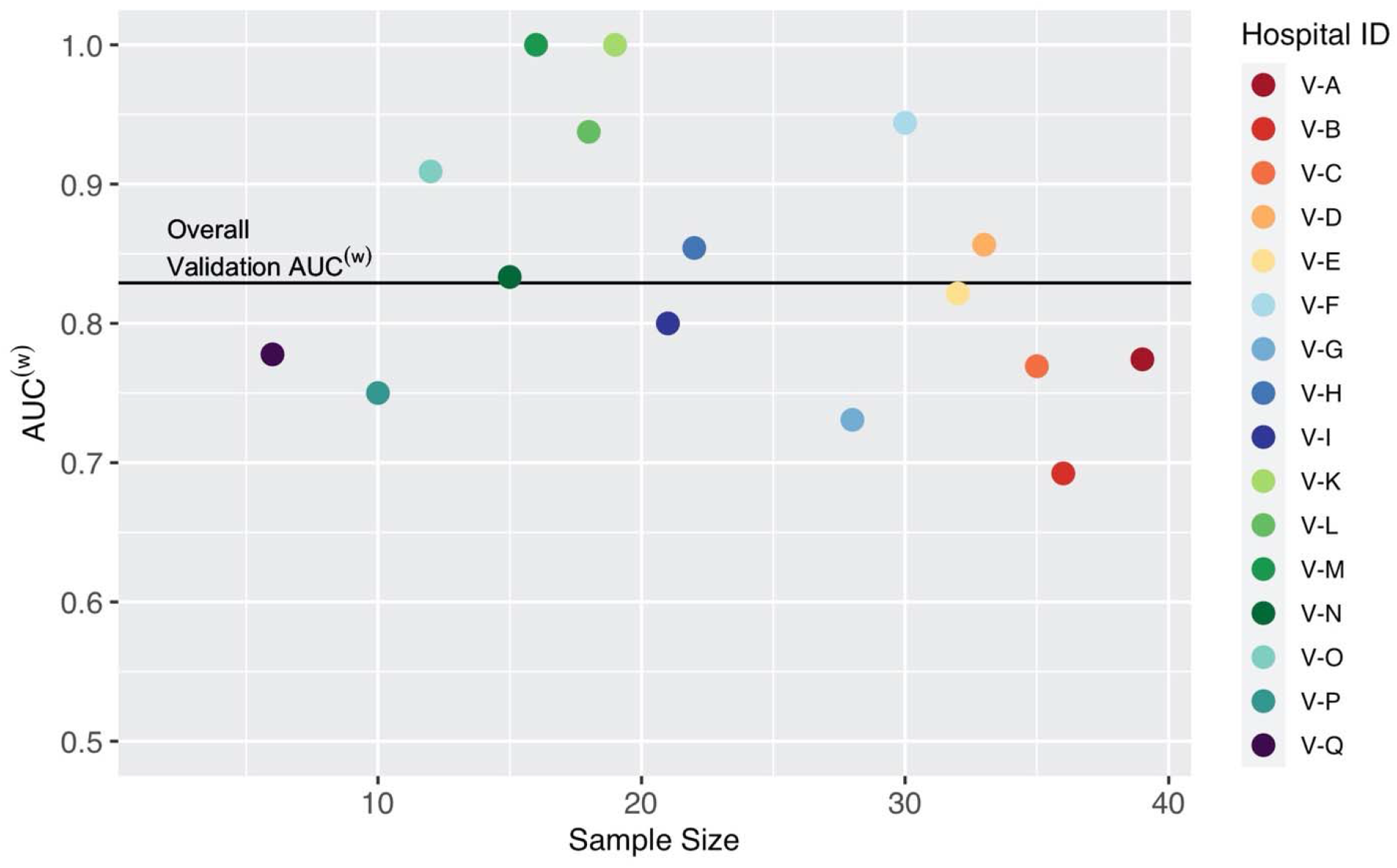
AUC^(w)^ by hospital in the validation set with overall validation AUC^(w)^. Hospitals V-J, V-R, and V-S are not included because they have observed COVID-19 mortality rates of 0% or 100%.

**Figure 2:**
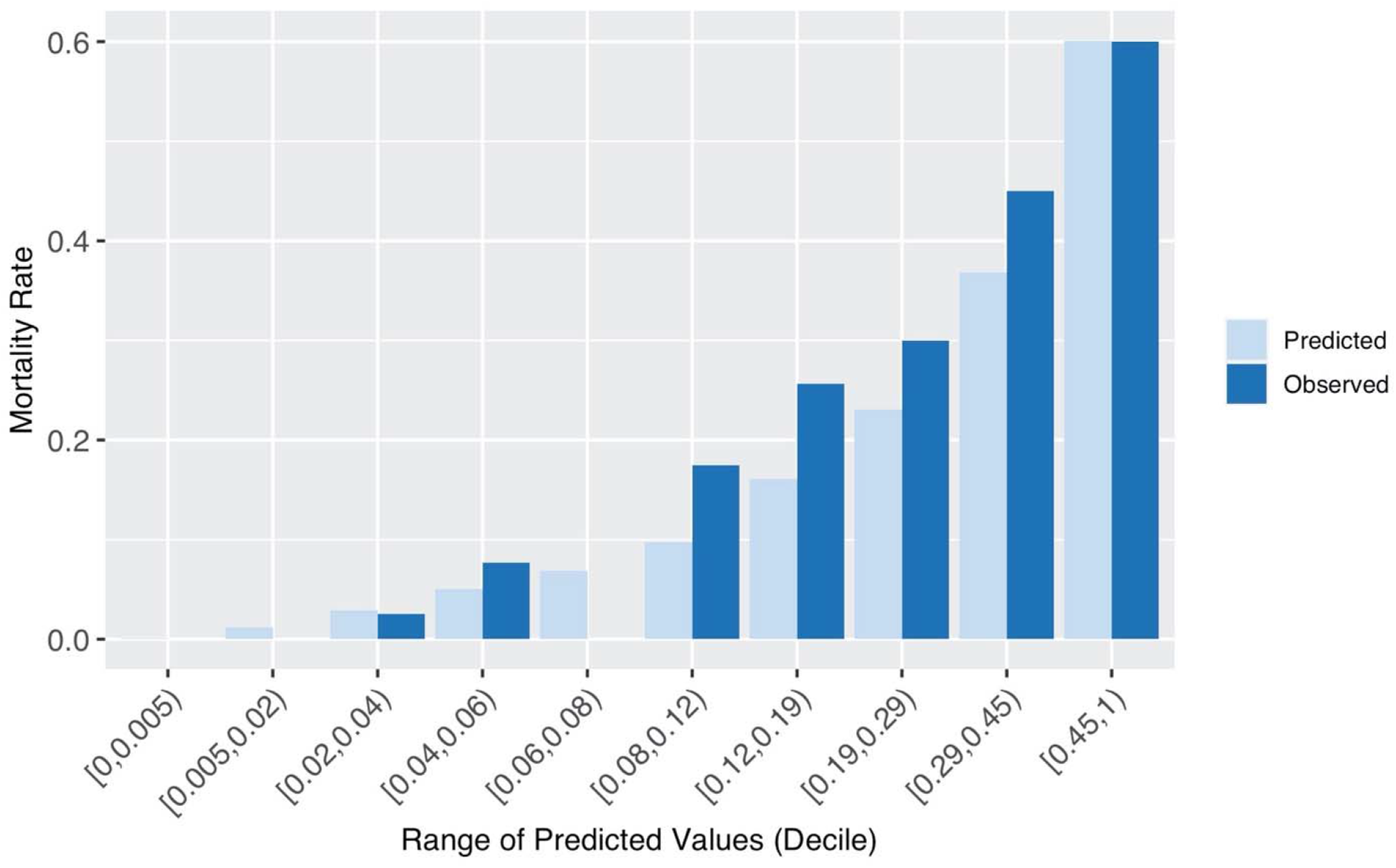
**Observed COVID-19 mortality rate and average predicted risk of COVID-19 mortality within each decile of predictions for the validation set.** Predictions use the individual hospital mortality rate.

## 4. Discussion

Using a systematic variable selection approach, we built a simple model that estimates the risk of in-hospital mortality for patients hospitalized for COVID-19 with comparable discrimination to more complex models. We performed external validation of the model in 19 separate hospitals for our validation set. It is notable that only age and a few vital signs and labs on hospital presentation and the hospital specific COVID-19 mortality rate are able to predict the risk of in-hospital mortality with high discrimination. Further, our model can be used even when not all variables are available to the user.

Age, creatinine, respiratory rate, and pulse oximetry have been found to be associated with mortality in COVID-19 patients in other studies with different populations, attesting to their validity as outcome predictors^2–5, 7^. Furthermore, the model was developed and validated on data from 39 diverse hospitals from different hospital systems, with variable size, urbanicity, profit status, and academic affiliation. Because of the diversity of hospitals included in Mi-COVID19, we expect that our risk score will generalize well to other hospitals, including those outside the state of Michigan.

We diverge from previous models by including individual hospital’s mortality rate for patients with COVID-19 in our model. The inclusion of this variable controls for unmeasured differences between hospitals, as well as the patient populations. We expect including this indicator for each hospital helps control for differing social determinants of health. We do find that after conditioning on the hospital, a patient’s race is not predictive of in-hospital mortality. If a user is uncertain about the current mortality rate at their hospital, they can still calculate a risk of mortality for a patient using our web application, as the mean COVID-19 mortality rate in the data (20%) is used automatically. Additionally, a user can compare the risk of mortality between patients within the same hospital without needing to know a hospital COVID-19 mortality rate. The contribution of the mortality rate to the prediction of mortality risk for each patient can be thought of as adjusting the constant term in the model for each hospital, based on previous understanding of the overall risk of COVID-19 mortality at that hospital. In other words, this is essentially incorporating a pretest probability into the model, which can be updated over time as the mortality rate at an individual hospital changes.

Our study should be interpreted in the context of some limitations. Many of the hospitalizations in our dataset were in Southeastern Michigan during the spring 2020 COVID-19 surge, when many hospitals in this region were experiencing very high patient volumes and treatment differed from current best practices. For example, in March and April 2020, dexamethasone and remdesivir were used only rarely, while hydroxychloroquine use was common. Thus, because our model was developed and validated using data from the Spring 2020 surge, it may overestimate the in-hospital mortality of patients treated in non-surge settings and with current best practices. Importantly, however, our model includes the hospital’s mortality rate for COVID-19 as a predictor, such that the model automatically re-calibrates over time. Furthermore, while in-hospital mortality has changed over time in relation to patient volume and the introduction of new therapies, we expect that age, respiratory rate, pulse oximetry, and creatinine will remain important predictors of in-hospital mortality for COVID-19, as these variables are consistently identified for inclusion in risk-prediction models. However, future studies that evaluate the model discrimination and calibration for patients hospitalized after the summer of 2020 will need to confirm that the model performance does not degrade over time.

In sum, we developed a parsimonious risk-prediction model for in-hospital mortality in patients from COVID-19. The use of data from a statewide registry, systematic approach to variable inclusion, and external validation should improve applicability in diverse hospital settings.

## Data Availability

The Mi-COVID19 data registry is proprietary. The data may be requested from the Michigan Hospital Medicine Safety Consortium.

https://www.mi-hms.org/quality-initiatives/mi-covid19-initiative

## Acknowledgements

We thank all BCBSM Collaborative Quality Initiatives that partnered together on data collection and all hospitals that volunteered to be part of Mi-COVID19. CZM and JAGB report support from the US National Science Foundation grant DMS-1646108. All authors report support from Blue Cross Blue Shield of Michigan and Blue Care Network. To our best knowledge, we have no other conflicts of interest to report.

## Notes

### Author Declarations

The study was deemed exempt by the University of Michigan Institutional Review Board.

