## Supplementary Material for "Derivation and external validation of a simple risk score to predict in-hospital mortality in patients hospitalized for COVID-19"

**eMethods.** Statistical methods supplement

**eTable 1.** List of predictive factors in the Mi-COVID19 data registry considered.
**eTable 2.** Characteristics of hospitals in the Mi-COVID19 data registry.
**eTable 3.** Full data patient characteristics, by derivation and validation set.

**eFigure 1.** Web application user interface.
**eFigure 2.** Mortality rate by hospital in the Mi-COVID19 data registry.
**eFigure 4.** Odds ratios and 95% confidence intervals for age in final risk model (reference age 50 years).

**eAppendix A.** Model selection details

- **eFigure 3a.** Change in quality metrics for top 10 variables with most improved MSE when added to base model of in-hospital mortality from COVID-19 on hospital COVID-19 mortality rate.
- **eFigure 3b.** Change in quality metrics for top 10 variables with most improved MSE when added to base model of in-hospital mortality from COVID-19 on patient's age and hospital COVID-19 mortality rate.
- **eFigure 3c.** Change in quality metrics for top 10 variables with most improved MSE when added to base model of in-hospital mortality from COVID-19 on patient's age, respiratory rate on presentation, pulse oximetry on presentation, heart rate on presentation, and hospital COVID-19 mortality rate.
- **eFigure 3d.** Change in quality metrics for top 10 variables with most improved MSE when added to base model of in-hospital mortality from COVID-19 on patient's age, respiratory rate on presentation, pulse oximetry on presentation, heart rate on presentation, creatinine on presentation, and hospital COVID-19 mortality rate.
- **eFigure 3e.** Change in quality metrics for backward selection when removed from base model of in-hospital mortality from COVID-19 on patient's age, respiratory rate on presentation, creatinine on presentation, pulse oximetry on presentation, heart rate on presentation, and hospital COVID-19 mortality rate.
- **eFigure 3f.** Change in quality metrics for top 10 variables with most improved MSE when added to base model of in-hospital mortality from COVID-19 on patient's age, respiratory rate on presentation, pulse oximetry on presentation, creatinine on presentation, and hospital COVID-19 mortality rate.

**eAppendix B.** Model discrimination within subgroups

- **eTable 4:** AUC by subgroup.
  - (A) Race
  - (B) Gender
  - (C) Age

### eMethods: Statistical methods supplement

### Adjusted AUC calculation

The AUC, which is defined as the area under the ROC curve, may be alternatively formulated as an estimate of the probability that a model correctly predicts a greater risk of mortality for a patient who actually died than for a patient who actually survived. Let A denote a randomly selected patient, and let B denote a randomly selected patient with the opposite outcome (i.e., if patient A died, patient B survived and vice versa). Then, let $p_A$ and $p_B$ be the predicted probabilities of in-hospital mortality for patient A and patient B, respectively. We say that the set of predictions $(A,\ B,\ p_A\ ,p_B\ )$ are "correct" if the patient who died had a larger predicted probability of mortality, "tied" if the predicted probabilities are equal, and "incorrect" if the patient who survived had a larger predicted probability of mortality. Then, the AUC is the average of the function $\psi(A,\ B,\ p_A\ ,p_B\ )$ defined below across all possible randomly selected pairs of predictions.

$$\psi(A,\ B,\ p_A\ ,p_B\ ) = \begin{cases} 1 \ \ if \text{correct} \\ \frac{1}{2} \qquad if \text{ tied} \\ 0 \ if \text{ incorrect} \end{cases}.$$

In the setting where the data includes patients from multiple hospitals and the mortality rate within each hospital is included in the model, the predicted probabilities for patients in a hospital with a higher mortality rate will already be larger than the predicted probabilities for patients in a hospital with a lower mortality rate. Therefore, there is built in discrimination between patients from different hospitals that has nothing to do with the clinical characteristics in the model. We only want to estimate the discrimination of the model based on the clinical characteristics included, without this artificial boost from including the hospital mortality rate, so we calculate an adjusted AUC instead, described below.

The key idea is that we modify the procedure described above so that we only compare pairs of patients (A, B) that are *in the same hospital*; as a result, any systematic differences between hospitals are irrelevant to the calculation of the adjusted AUC. More specifically, assume we have *m* hospitals and let $N_i$ denote the number of patients who are in hospital *i* and *N* denote the total number of patients across all *m* hospitals. As before, let A denote a randomly selected patient. However, now let B denote a randomly selected patient *from the same hospital* as patient A where patient B has the opposite outcome. We keep all other notation the same as above.

We can now calculate $AUC^{(w)}$ as:

$$\begin{aligned} AUC^{(w)} &= \mathrm{E}[\psi(A,\ B,\ p_A\ ,p_B\ )] \\ &= \textstyle\sum_{i=1}^{m} E[\psi(A,\ B,\ p_A\ ,p_B\ )|\text{A in hospital i}]\mathrm{P}(\text{A in hospital } i) \\ &= \textstyle\sum_{i=1}^{m} E[\psi(A,\ B,\ p_A\ ,p_B\ )|\text{A and B in hospital i}]\mathrm{P}(\text{A in hospital } i) \\ &= \textstyle\sum_{i=1}^{m} E[\psi(A,\ B,\ p_A\ ,p_B\ )|\text{A and B in hospital } i]\frac{N_i}{N} \\ &= \textstyle\sum_{i=1}^{m} \frac{N_i}{N} AUC_i \end{aligned}$$

Where $AUC_i$ denotes the within-hospital AUC for hospital i, i.e.,

$$\text{AUC}_i = E[\psi(A,\ B,\ p_A\ , p_B\ )|\text{A and B in hospital } i]$$

So, $\text{AUC}^{(w)}$ can be written as:

$$\text{AUC}^{(w)} = \sum_{i=1}^{m} w_i\, AUC_i$$

$$w_i = \frac{N_i}{N}.$$

Therefore, $\text{AUC}^{(w)}$ is a weighted average of the individual hospital AUC's with weights proportional to the hospital sample size.

### Modeling for web application

We use the following technique to allow for approximate refitting of the model without sharing the proprietary dataset when we share the model in the web application. First, we fit the logistic regression model described on the mortality outcome. Next, we transformed the predictions using a logit transform. If you fit a linear regression model (OLS) on these transformed predictions, the coefficients of the OLS model are exactly the same as those in the original logistic regression model. The coefficients of an OLS model ($\beta$) can be calculated using matrix algebra with the design matrix (*X*) and the outcome vector (*y*):

$$\beta = (X'X)^{-1}(X'y)$$

We save the matrices *G = X'X and B = X'y* where *y* is the nx1 vector of logit transformed predictions from the logistic regression model and *X* is the design matrix.

Then, say that one wishes to estimate the coefficients for a model without creatinine. Let creatinine be the fourth column in *X* and let $G^*$ be *G* with the 4th row and column removed and $B^*$ be B with the 4th row removed. Then, we can estimate the coefficients $\beta^*$for a model without creatinine as:

$$\beta^* = (G^*)^{-1}(B^*)$$

Thus, we can estimate the coefficients of the OLS model with any subset of the variables that we included in our final model. This method allows us to still share a model that can be refitted and updated while maintaining data privacy since we save and share the *G* and *B* matrices rather than the raw data.

### Odds ratio and the hospital mortality rate

Note that the estimated odds ratio of mortality in a hospital with mortality rate $r$, for a patient with other covariates $X_1$ as compared to a patient with other covariates $X_2$, where $\gamma$ is the model coefficient for $r$and $\beta$ are the coefficients for all other covariates is $e^{\gamma r+\ \beta' X_1}/e^{\gamma r+\beta' X_2} = e^{\beta' X_1-\beta' X_2}$. The hospital mortality rate is cancelled out in the odds ratio. Therefore, the estimated odds ratio between two patients in the same hospital can be calculated without knowing the hospital mortality rate.

| Category | Variable |
|---|---|
| Demographics and admission | Age |
| | Gender |
| | Race |
| | Ethnicity |
| | Admission reason |
| | Prior residence |
| | How did the patient arrive to the ED? |
| | BMI |
| History | Smoking history |
| | Vaping history |
| | Left ventricular ejection fraction documented prior to admission? |
| | Does the patient have a history of Aortic Stenosis? |
| | Has the patient received any type of dialysis prior to the hospital encounter? |
| | Has the patient been on a home ventilator prior to the hospital encounter? |
| | Is the patient on home oxygen? |
| | Is the patient a healthcare worker? |
| | Is the patient a service worker? |
| | Previously treated by an opioid |
| | Previously treated by a benzodiazepine |
| | Previously treated by a sedative |
| Comorbidities | AIDS or HIV |
| | Diabetes - Uncomplicated |
| | Hypertension |
| | Hemiplegia or Paraplegia |
| | Inflammatory Bowel Disease (i.e., Crohns or Ulcerative Colitis) |
| | Leukemia |
| | Lymphoma |
| | Any Malignancy without Metastasis |
| | Metastatic Solid Tumor |
| | Mild Liver Disease |
| | Moderate or Severe Liver Disease |
| | Asthma |
| | Moderate or Severe Kidney Disease |
| | Myocardial Infarction (MI) (history of/prior event) |
| | Transplant |
| | Peptic Ulcer Disease |
| | Peripheral Vascular Disorders |
| | Rheumatoid Arthritis or Arthropathy/Connective Tissue |
| | Venous Thromboembolism (DVT/PE) |
| | Cardiovascular Disease |
| | Cerebrovascular Disease |
| | Chronic obstructive pulmonary disease (COPD) |
| | Congestive heart failure (CHF)/Cardiomyopathy |
| | Chronic Pulmonary Disease (other than asthma or COPD) |
| | Dementia |
| | Diabetes - Complicated |
| | Number of comorbidities |
| Symptoms | Fever (measured temperature 99.0 - 100.4 [F]) |
| | Fatigue |
| | Diarrhea |
| | Nausea/vomiting |
| | Altered Mental Status |

| Category | Variable |
|---|---|
| Symptoms cont. | None of the above |
| | Fever (measured temperature >100.4 [F]) |
| | Subjective fever |
| | Generalized malaise |
| | Weakness |
| | Loss of taste |
| | Dyspnea / shortness of breath |
| | Loss of smell |
| | Hypoxia / new or escalated O2 requirement |
| | Cough (New or Worsening) |
| | Non-pleuritic chest pain |
| | Pleuritic chest pain |
| | Sputum productions |
| | Rhinorrhea |
| | Myalgias |
| Chief Complaint | Difficulty breathing or shortness of breath |
| | Cough |
| | Fever |
| | Chest pain |
| | Nausea or vomiting |
| | Diarrhea |
| | Other chief complaint |
| First recorded vitals during the hospital encounter (vital signs from ED if admitted from the ED) | Temperature |
| | Heart rate |
| | Respiratory rate |
| | Systolic blood pressure |
| | Diastolic blood pressure |
| | Pulse oximetry |
| | Triage score |
| Labs from day 1 or day 2 of hospitalization (first available) | Highest Lactate |
| | Highest Creatinine |
| | Highest Alanine Amino Transferase (ALT) |
| | Highest Total Bilirubin |
| | Highest White Blood Cell (WBC) |
| | Highest Troponin |
| | Highest Brain Natriuretic Peptide (BNP) |
| | Highest Ferritin |
| | Highest C-reactive protein (CRP) |
| | Highest Lactic Acid Dehydrogenase (LDH) |
| | Highest Procalcitonin |
| | Highest Hemoglobin (Hgb) |
| | Lowest Platelet |
| | Lowest Absolute Lymphocyte Count |
| | Lowest pH |
| | Highest Fibrinogen |
| | Highest Interleukin 6 (IL-6) |
| | Highest erythrocyte sedimentation rate (ESR) |
| Chest x-ray results from day 1 or day 2 of hospitalization (first available) | Air Spaced Density Disease |
| | Loculations |
| | New or Worsening Infiltrates |
| | Nodular Airspace Disease |
| | Mass |

| Category | Variable |
|---|---|
| Chest x-ray results from day 1 or day 2 of hospitalization (first available) | Pleural Effusion |
| | Pneumonia |
| | Pulmonary Edema |
| | Pulmonary Vascular Congestion |
| | No Evidence of Pneumonia |
| | No Change from Previous/No Interval Change |
| | Atelectasis |
| | Normal/No Abnormalities |
| | None of the Above Statements |
| | Post Obstructive Pneumonia |
| | Necrotizing Pneumonia |
| | Nodules |
| | Aspiration |
| | Aspiration Pneumonia |
| | Bronchial Wall Thickening/Pleural Thickening |
| | Bronchiectasis |
| | Emphysema/Emphysematous Changes |
| | Bronchopnuemonia |
| | Hyperinflation |
| | Infection (Cannot Rule Out Infection, Likely Infection) |
| | Infiltrate (Not Specified) |
| | Interstitial Lung Disease/Interstitial Disease |
| | Neoplasm/Metastatic Disease/Malignancy |
| | Mucus Plugging/Plugging |
| | Pneumonitis |
| | Tree in Bud |
| | Interval Improvement or Resolution |
| | Abscess |
| | Cannot Rule Out Pneumonia |
| | Granuloma |
| | Opacities (central) |
| | Opacities (peripheral) |
| | Opacities (subpleural) |
| | Opacities (Not specified) |
| | Cavitation |
| | Consolidation |
| | Ground Glass |
| | Infiltrate (Single Lobe) |
| | Infiltrate (Multiple Lobes) |
| | Other abnormal finding |
| | Any pneumonia indication on chest x-ray |

**eTable 1: List of predictive factors in the Mi-COVID19 data registry considered.**

| | Data Set Specific Characteristics | | | | General Characteristics | | | |
|---|---|---|---|---|---|---|---|---|
| Hospital ID | Full Sample Size | Complete Cases Size | COVID Mortality Rate | Discharge Date Range in Data | Bed Size | Discharges | Teaching Hospital? | Hospital Type |
| A | 269 | 260 | 0.15 | 3/17 - 5/26 | 1,059 | 44,920 | Yes | Voluntary non-profit - Private |
| B | 155 | 142 | 0.21 | 3/18 - 5/20 | 537 | 30,614 | Yes | Voluntary non-profit - Church |
| C | 110 | 109 | 0.13 | 3/28 - 7/20 | 109 | 32,636 | Yes | Voluntary non-profit - Other |
| D | 108 | 102 | 0.14 | 3/15 - 5/15 | 1,070 | 61,758 | Yes | Voluntary non-profit - Private |
| E | 96 | 91 | 0.17 | 4/04 - 8/14 | 196 | 9,307 | No | Voluntary non-profit - Private |
| F | 90 | 89 | 0.11 | 3/13 - 6/21 | 283 | 15,855 | Yes | Voluntary non-profit - Church |
| G | 91 | 88 | 0.11 | 3/21 - 6/06 | 317 | 15,093 | Yes | Voluntary non-profit - Private |
| H | 88 | 85 | 0.49 | 3/21 - 4/10 | 404 | 18,345 | Yes | Proprietary |
| I | 83 | 79 | 0.28 | 3/19 - 6/01 | 443 | 19,102 | Yes | Voluntary non-profit - Private |
| J | 82 | 78 | 0.11 | 3/20 - 5/03 | 443 | 17,240 | Yes | Voluntary non-profit - Other |
| K | 72 | 71 | 0.08 | 3/18 - 5/04 | 250 | 12,186 | Yes | Voluntary non-profit - Private |
| L | 69 | 68 | 0.25 | 3/16 - 5/14 | 632 | 30,354 | Yes | Voluntary non-profit - Private |
| M | 67 | 65 | 0.15 | 3/16 - 4/30 | 330 | 13,159 | Yes | Voluntary non-profit - Private |
| N | 66 | 65 | 0.12 | 3/19 - 5/09 | 458 | 34,863 | Yes | Voluntary non-profit - Private |
| O | 62 | 61 | 0.19 | 3/23 - 5/01 | 158 | 7,704 | Yes | Proprietary |
| P | 67 | 58 | 0.37 | 3/18 - 5/21 | 304 | 15,804 | Yes | Voluntary non-profit - Private |
| Q | 54 | 48 | 0.07 | 3/16 - 6/23 | 189 | 8,639 | No | Voluntary non-profit - Private |
| R | 45 | 45 | 0.31 | 3/22 - 6/04 | 378 | 17,969 | Yes | Voluntary non-profit - Private |
| S | 49 | 42 | 0.27 | 3/22 - 5/09 | 434 | 26,705 | Yes | Voluntary non-profit - Private |
| T | 46 | 44 | 0.54 | 3/23 - 4/19 | 273 | 10,815 | Yes | Proprietary |
| V-A | 39 | 39 | 0.21 | 3/18 - 7/07 | 391 | 21,759 | Yes | Voluntary non-profit - Other |
| V-B | 39 | 36 | 0.33 | 3/17 - 4/22 | 584 | 19,882 | Yes | Proprietary |
| V-C | 37 | 35 | 0.24 | 3/20 - 4/14 | 215 | 7,797 | Yes | Voluntary non-profit - Private |
| V-D | 37 | 33 | 0.27 | 3/16 - 4/05 | 877 | 35,908 | Yes | Voluntary non-profit - Private |
| V-E | 33 | 32 | 0.12 | 3/17 - 4/22 | 189 | 6,142 | Yes | Voluntary non-profit - Private |
| V-F | 30 | 30 | 0.17 | 3/21 - 5/10 | 193 | 9,816 | Yes | Voluntary non-profit - Private |
| V-G | 29 | 28 | 0.07 | 4/02 - 5/22 | 208 | 10,476 | Yes | Voluntary non-profit - Private |
| V-H | 23 | 22 | 0.26 | 3/16 - 4/12 | 361 | 18,166 | Yes | Voluntary non-profit - Private |

| | | | | | | | | |
|---|---|---|---|---|---|---|---|---|
| V-I | 21 | 21 | 0.24 | 3/22 - 6/03 | 136 | 2,767 | Yes | Voluntary non-profit - Other |
| V-J | 22 | 20 | 0.00 | 3/14 - 4/12 | 191 | 12,260 | Yes | Voluntary non-profit - Private |
| V-K | 20 | 19 | 0.05 | 3/05 - 5/11 | 133 | 3,763 | No | Voluntary non-profit - Private |
| V-L | 19 | 18 | 0.16 | 3/27 - 5/02 | 179 | 7,254 | No | Proprietary |
| V-M | 16 | 16 | 0.06 | 4/06 - 6/12 | 79 | 3,349 | Yes | Voluntary non-profit - Private |
| V-N | 18 | 15 | 0.22 | 3/23 - 5/26 | 360 | 14,206 | Yes | Voluntary non-profit - Private |
| V-O | 13 | 12 | 0.08 | 4/01 - 4/20 | 186 | 11,579 | Yes | Voluntary non-profit - Private |
| V-P | 12 | 10 | 0.17 | 4/05 - 6/11 | 328 | 15,767 | Yes | Voluntary non-profit - Other |
| V-Q | 7 | 6 | 0.43 | 4/14 - 6/17 | 139 | 4,362 | No | Proprietary |
| V-R | 7 | 5 | 0.00 | 3/13 - 4/17 | 310 | 11,233 | Yes | Voluntary non-profit - Private |
| V-S | 1 | 1 | 1.00 | 4/14 - 4/14 | 78 | 4,953 | No | Voluntary non-profit - Other |
| V-T | 1 | 0 | 1.00 | 3/28 - 3/28 | 365 | 18,252 | Yes | Voluntary non-profit – Other |

Hospitals are ordered by the complete cases sample size for the final risk score model used. “Full Sample” is the sample size available before removing observations with missing data for the variables in the final model. The mortality rate is calculated using the full sample. A “V” at the beginning of a hospital ID indicates that that hospital was in the validation set. Hospital V-T was not included in the final validation set because there were no complete cases for that hospital.

#### eTable 2: Characteristics of hospitals in the Mi-COVID19 data registry.

| Characteristic | Overall [N = 2193] mean/No. (SD/%) [n*] | Dataset Derivation [N = 1769] | Validation [N = 424] |
|---|---|---|---|
| Age | 63.9 (16.8) [2193] | 64.4 (16.7) [1769] | 61.5 (17.3) [424] |
| Gender (female) | 1046 (48%) [2193] | 830 (47%) [1769] | 216 (51%) [424] |
| Race (yes) | | | |
| Black | 1024 (49%) [2101] | 840 (49%) [1698] | 184 (46%) [403] |
| White | 949 (45%) [2101] | 744 (44%) [1698] | 205 (51%) [403] |
| Asian | 52 (2%) [2101] | 45 (3%) [1698] | 7 (2%) [403] |
| Native American or Pacific Islander | 10 (0%) [2101] | 10 (1%) [1698] | 0 (0%) [403] |
| Other | 66 (3%) [2101] | 59 (3%) [1698] | 7 (2%) [403] |
| Ethnicity (yes) | | | |
| Hispanic | 113 (5%) [2185] | 92 (5%) [1762] | 21 (5%) [423] |
| Non-Hispanic | 1907 (87%) [2185] | 1535 (87%) [1762] | 372 (88%) [423] |
| Unknown | 165 (8%) [2185] | 135 (8%) [1762] | 30 (7%) [423] |
| Residing in a Nursing Facility or Assisted Living (yes) | 408 (19%) [2164] | 340 (19%) [1748] | 68 (16%) [416] |
| Ever-smoker (yes) | 809 (40%) [2041] | 645 (39%) [1648] | 164 (42%) [393] |
| BMI | 32.6 (60.4) [2079] | 31.2 (8.5) [1680] | 38.7 (136.7) [399] |
| No. of comorbidities | | | |
| 0 | 289 (13%) [2187] | 228 (13%) [1769] | 61 (15%) [418] |
| 1 | 420 (19%) [2187] | 341 (19%) [1769] | 79 (19%) [418] |
| 2 | 462 (21%) [2187] | 369 (21%) [1769] | 93 (22%) [418] |
| 3 | 375 (17%) [2187] | 299 (17%) [1769] | 76 (18%) [418] |
| 4 | 269 (12%) [2187] | 225 (13%) [1769] | 44 (11%) [418] |
| >4 | 372 (17%) [2187] | 307 (17%) [1769] | 65 (16%) [418] |
| Presence of comorbidity (yes) | | | |
| Cardiovascular disease | 596 (27%) [2187] | 485 (27%) [1769] | 111 (27%) [418] |
| Congestive heart failure | 334 (15%) [2187] | 275 (16%) [1769] | 59 (14%) [418] |
| Chronic obstructive pulmonary disease | 273 (12%) [2187] | 208 (12%) [1769] | 65 (16%) [418] |
| Asthma | 274 (13%) [2187] | 219 (12%) [1769] | 55 (13%) [418] |
| Diabetes (complicated and uncomplicated) | 817 (37%) [2187] | 655 (37%) [1769] | 162 (39%) [418] |
| Severe liver disease | 15 (1%) [2187] | 12 (1%) [1769] | 3 (1%) [418] |
| Cancer | 175 (8%) [2187] | 144 (8%) [1769] | 31 (7%) [418] |
| Symptoms (yes) | | | |
| Fatigue | 747 (34%) [2187] | 585 (33%) [1769] | 162 (39%) [418] |
| Fever (subjective and objective) | 1827 (84%) [2187] | 1452 (82%) [1769] | 375 (90%) [418] |
| Chest pain | 358 (16%) [2187] | 301 (17%) [1769] | 57 (14%) [418] |
| Hypoxia | 878 (40%) [2187] | 729 (41%) [1769] | 149 (36%) [418] |
| First recorded heart rate | | | |
| < 90 BPM | 847 (39%) [2177] | 701 (40%) [1760] | 146 (35%) [417] |
| 90-100 BPM | 495 (23%) [2177] | 391 (22%) [1760] | 104 (25%) [417] |
| 101-124 BPM | 683 (31%) [2177] | 544 (31%) [1760] | 139 (33%) [417] |
| > 124 BPM | 152 (7%) [2177] | 124 (7%) [1760] | 28 (7%) [417] |
| First recorded respiratory rate | | | |
| < 20 | 809 (38%) [2149] | 645 (37%) [1734] | 164 (40%) [415] |
| 20-24 | 855 (40%) [2149] | 682 (39%) [1734] | 173 (42%) [415] |
| 25-30 | 284 (13%) [2149] | 240 (14%) [1734] | 44 (11%) [415] |
| > 30 | 201 (9%) [2149] | 167 (10%) [1734] | 34 (8%) [415] |

| Characteristic | Overall [N = 2193] mean/No. (SD/%) [n*] | Dataset Derivation [N = 1769] | Validation [N = 424] |
|---|---|---|---|
| First recorded systolic blood pressure | | | |
| >= 101 mmHg | 1997 (93%) [2154] | 1619 (93%) [1740] | 378 (91%) [414] |
| 90 - 100 mmHg | 101 (5%) [2154] | 80 (5%) [1740] | 21 (5%) [414] |
| < 90 mmHg | 56 (3%) [2154] | 41 (2%) [1740] | 15 (4%) [414] |
| First recorded pulse oximetry | | | |
| 91-100% | 1726 (80%) [2164] | 1386 (79%) [1750] | 340 (82%) [414] |
| 81-90% | 342 (16%) [2164] | 287 (16%) [1750] | 55 (13%) [414] |
| 71-80% | 56 (3%) [2164] | 45 (3%) [1750] | 11 (3%) [414] |
| <= 70% | 40 (2%) [2164] | 32 (2%) [1750] | 8 (2%) [414] |
| Triage score | | | |
| 1 | 112 (6%) [1880] | 91 (6%) [1564] | 21 (7%) [316] |
| 2 | 907 (48%) [1880] | 731 (47%) [1564] | 176 (56%) [316] |
| 3 | 753 (40%) [1880] | 646 (41%) [1564] | 107 (34%) [316] |
| 4 | 47 (3%) [1880] | 37 (2%) [1564] | 10 (3%) [316] |
| 5 | 61 (3%) [1880] | 59 (4%) [1564] | 2 (1%) [316] |
| Highest initial creatinine (mg/dL) | 1.6 (1.7) [2140] | 1.7 (1.7) [1737] | 1.5 (1.7) [403] |
| Highest initial white blood cell count (K/uL) | 8.3 (6.4) [2154] | 8.4 (6.7) [1750] | 7.9 (4.4) [404] |
| Pneumonia indication on chest x-ray (yes) | 1618 (79%) [2054] | 1318 (78%) [1684] | 300 (81%) [370] |

* n is the number of complete cases in the data for the given variable. Percentages are calculated as No./n.

**eTable 3: Full data patient characteristics, by derivation and validation set.**

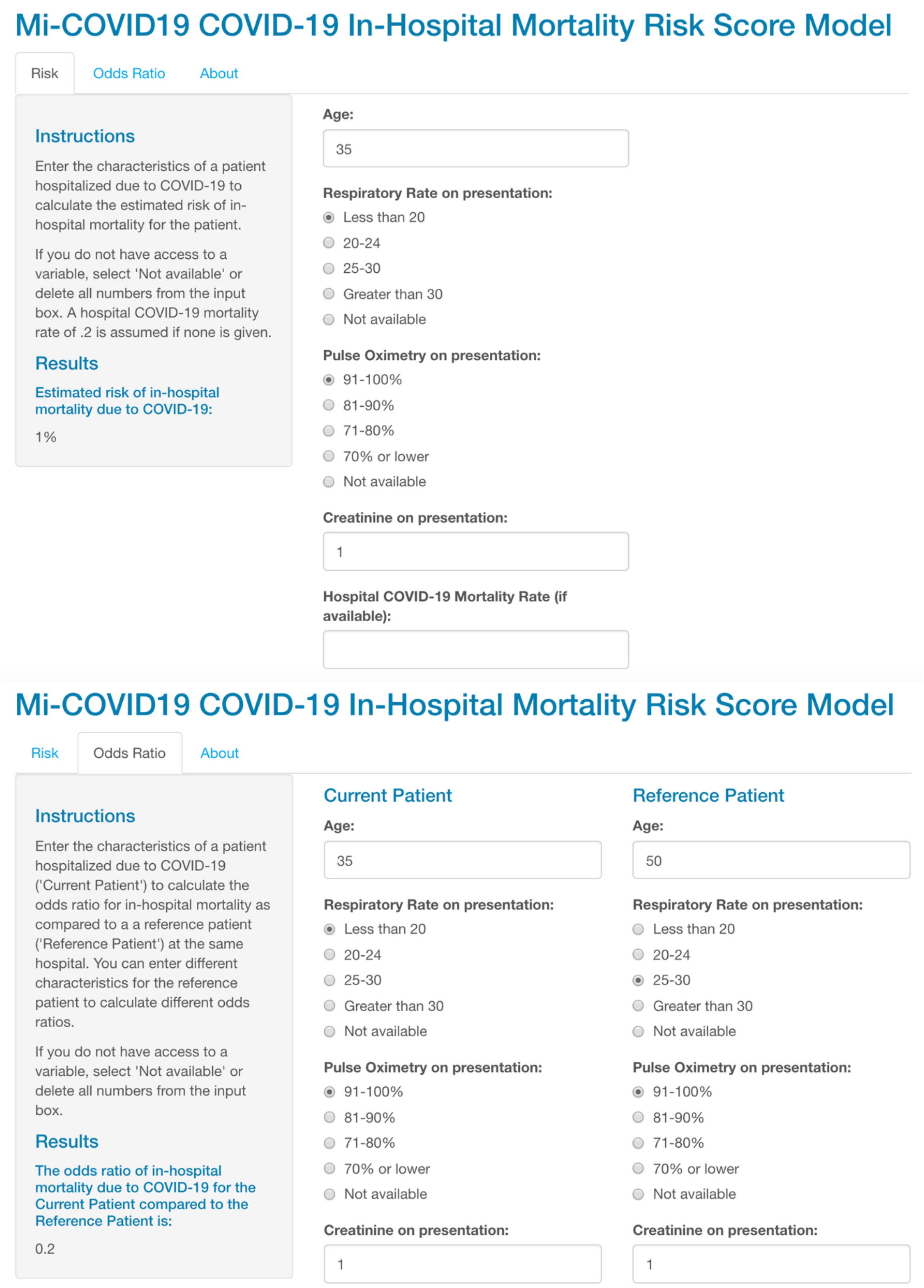


**eFigure 1: Web application user interface.** The app can be accessed at https://micovidriskcalc.org/.

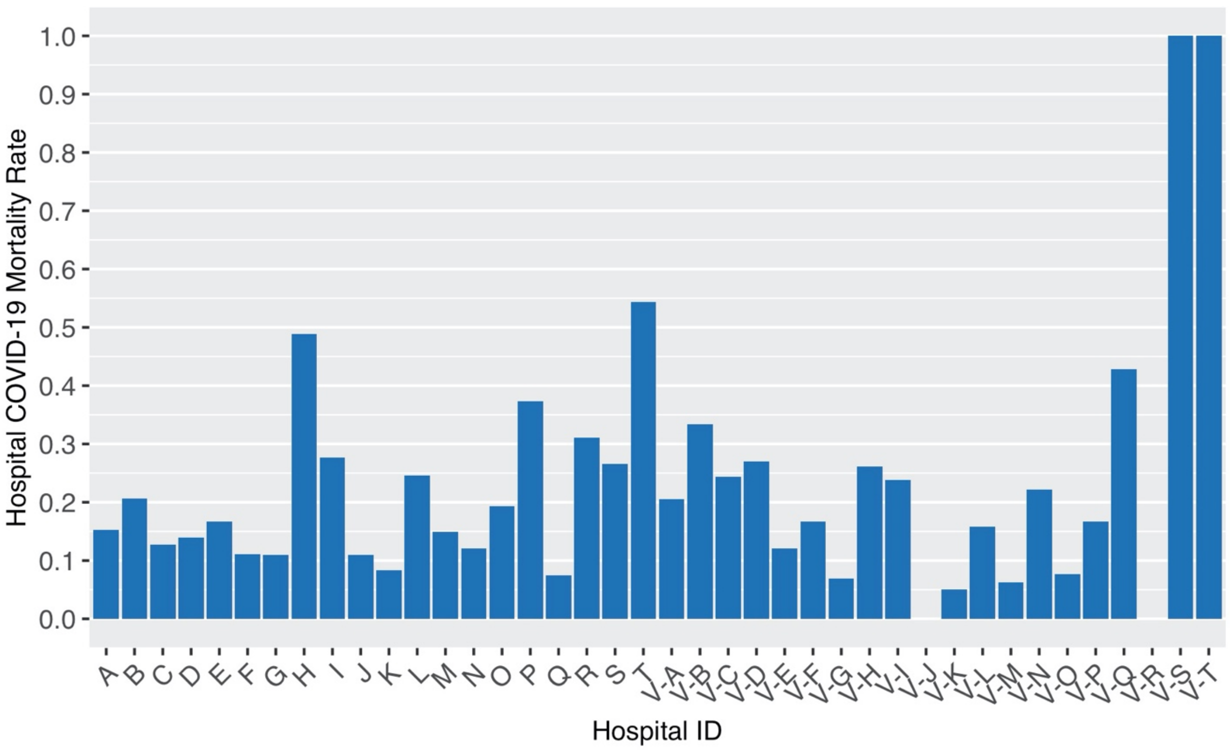


**eFigure 2: Mortality rate by hospital in the Mi-COVID19 data registry.** The mortality rate is calculated using the full sample of patients who tested positive for SARS-CoV-2 available in the Mi-COVID19 data. Hospitals are ordered by the complete cases sample size for the final risk model. A “V” at the beginning of a hospital ID indicates that that hospital was in the validation set. Hospital V-T was not included in the final validation set because there were no complete cases for that hospital.

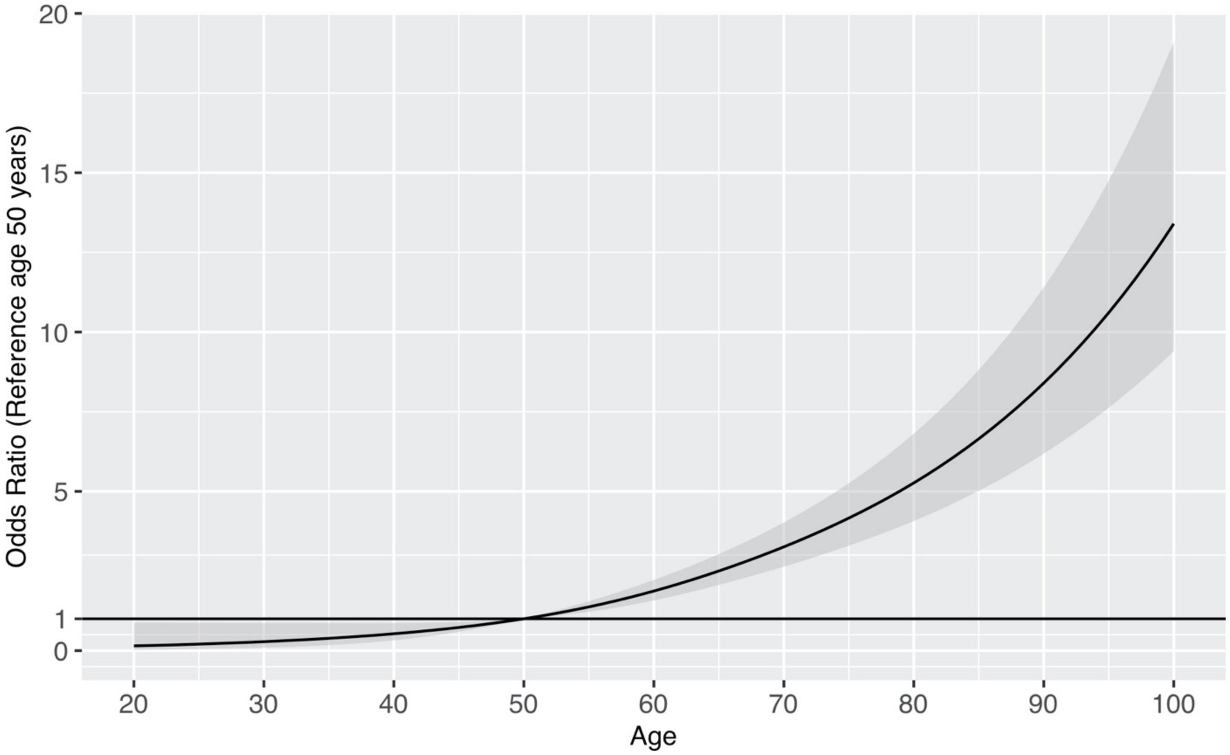


**eFigure 4. Odds ratio and 95% confidence interval for age in final risk model with a reference age of 50 years old.** The grey shaded region represents the 95% confidence interval for the odds ratio.

**eAppendix A: Model selection details**

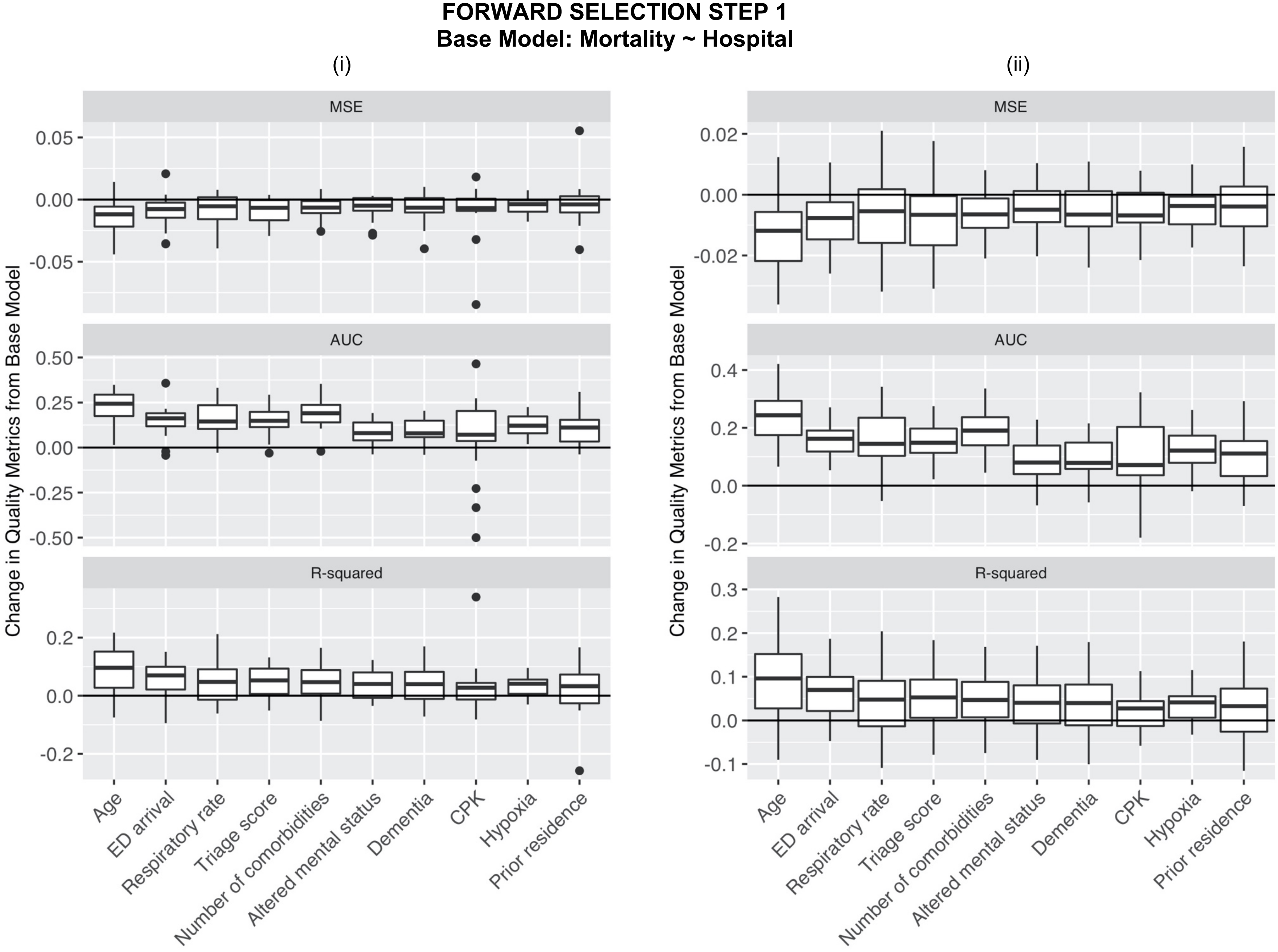

(iii)

| Variable | N | Change from Base Model<br>Mortality ~ Hospital<br>MSE | AUC$^{(w)}$ | R-squared |
|---|---|---|---|---|
| **Age** | **1769** | **-0.013** | **0.220** | **0.081** |
| ED arrival | 1659 | -0.010 | 0.156 | 0.063 |
| Respiratory rate | 1734 | -0.008 | 0.149 | 0.051 |
| Triage score | 1505 | -0.007 | 0.147 | 0.047 |
| Number of comorbidities | 1769 | -0.007 | 0.190 | 0.044 |
| Altered mental status | 1769 | -0.006 | 0.087 | 0.038 |
| Dementia | 1769 | -0.006 | 0.087 | 0.037 |
| Creatine Phosphokinase (CPK) | 447 | -0.006 | 0.093 | 0.029 |
| Hypoxia | 1769 | -0.005 | 0.127 | 0.031 |
| Prior residence | 1746 | -0.004 | 0.108 | 0.025 |

**eFigure 3a: Change in quality metrics for top 10 variables with most improved MSE when added to base model of in-hospital mortality from COVID-19 on hospital COVID-19 mortality rate.** (i) Histograms of change in MSE, AUC$^{(w)}$ and R-squared for all 20 hospitals in the derivation set. (ii) Histograms of change in MSE, AUC$^{(w)}$ and R-squared for all 20 hospitals in the derivation set with outliers removed. (iii) Change in quality metrics for all derivation hospitals combined. Bolded variables were chosen for inclusion in the next step of forward selection.

Based on these results, we included age in the model for the next step of forward selection because it was the variable with the most improved MSE, AUC$^{(w)}$, and R-squared overall (iii) and showed improvement for almost all hospitals on these metrics (i and ii).

**FORWARD SELECTION STEP 2**
**Base Model: Mortality ~ Age + Hospital**

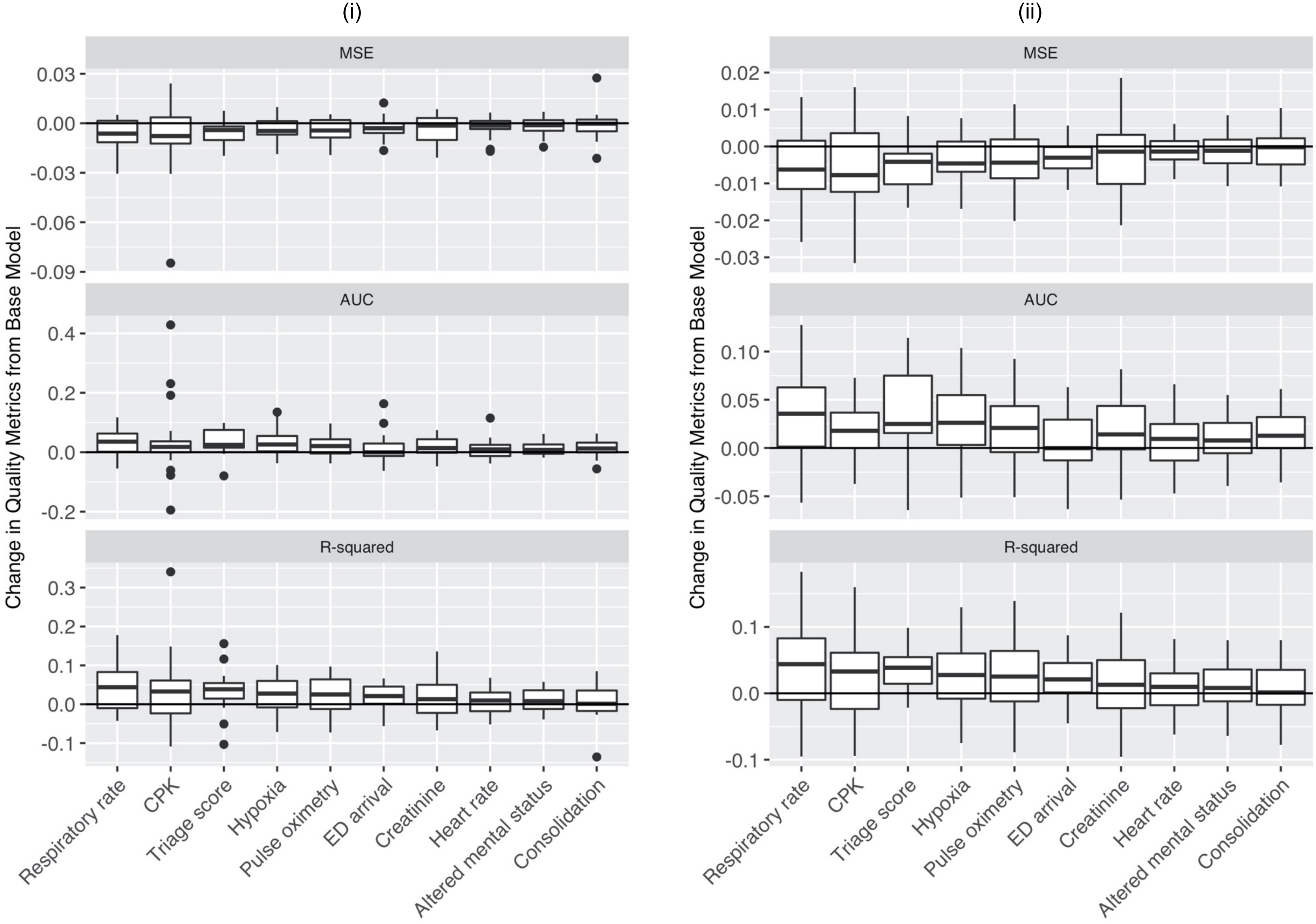

(iii)

| Variable | N | Change from Base Model Mortality ~ Age + Hospital MSE | AUC(w) | R-squared |
|---|---|---|---|---|
| **Respiratory rate** | **1734** | **-0.007** | **0.032** | **0.045** |
| Creatine Phosphokinase (CPK) | 447 | -0.006 | 0.041 | 0.031 |
| Triage score | 1505 | -0.006 | 0.033 | 0.037 |
| Hypoxia | 1769 | -0.004 | 0.035 | 0.028 |
| **Pulse oximetry** | **1750** | **-0.004** | **0.021** | **0.024** |
| ED arrival | 1659 | -0.003 | 0.012 | 0.019 |
| Highest creatinine | 1737 | -0.003 | 0.020 | 0.016 |
| **Heart rate** | **1760** | **-0.003** | **0.010** | **0.016** |
| Altered mental status | 1769 | -0.002 | 0.014 | 0.010 |
| Consolidation on chest x-ray | 1684 | -0.002 | 0.014 | 0.010 |

**eFigure 3b: Change in quality metrics for top 10 variables with most improved MSE when added to base model of in-hospital mortality from COVID-19 on patient's age and hospital COVID-19 mortality rate.** (i) Histograms of change in MSE, AUC(w) and R-squared for all 20 hospitals in the derivation set. (ii) Histograms of change in MSE, AUC(w) and R-squared for all 20 hospitals in the derivation set with outliers removed. (iii) Change in quality metrics for all derivation hospitals combined. Bolded variables were chosen for inclusion in the next step of forward selection.

Based on these results, we included respiratory rate, pulse oximetry, and heart rate in the model for the next step of forward selection. Respiratory rate was the variable with the most improved MSE and pulse oximetry and heart rate were two other vital signs that appeared to be predictive in the model overall (iii) and consistently across individual hospitals (i and ii). CPK was missing for a majority of patients and was unlikely to be widely available for COVID-19 patients. The triage score showed improvement, however, the other vital signs are included in the triage score. We also prioritized adding the vital signs over symptoms such as hypoxia. ED arrival described the manner in which a patient arrived at the hospital (such as "car," "ambulance," and "by foot"), which we did not expect to be widely available at all hospitals. Therefore, in the second step of forward selection, we decided to include the three most predictive vital signs. In the following steps, we could determine whether any of the other variables that appeared predictive in this step remained predictive after the vital signs were added to the model.

**FORWARD SELECTION STEP 3**
**Base Model: Mortality ~ Age + Respiratory Rate + Pulse Oximetry + Heart Rate + Hospital**

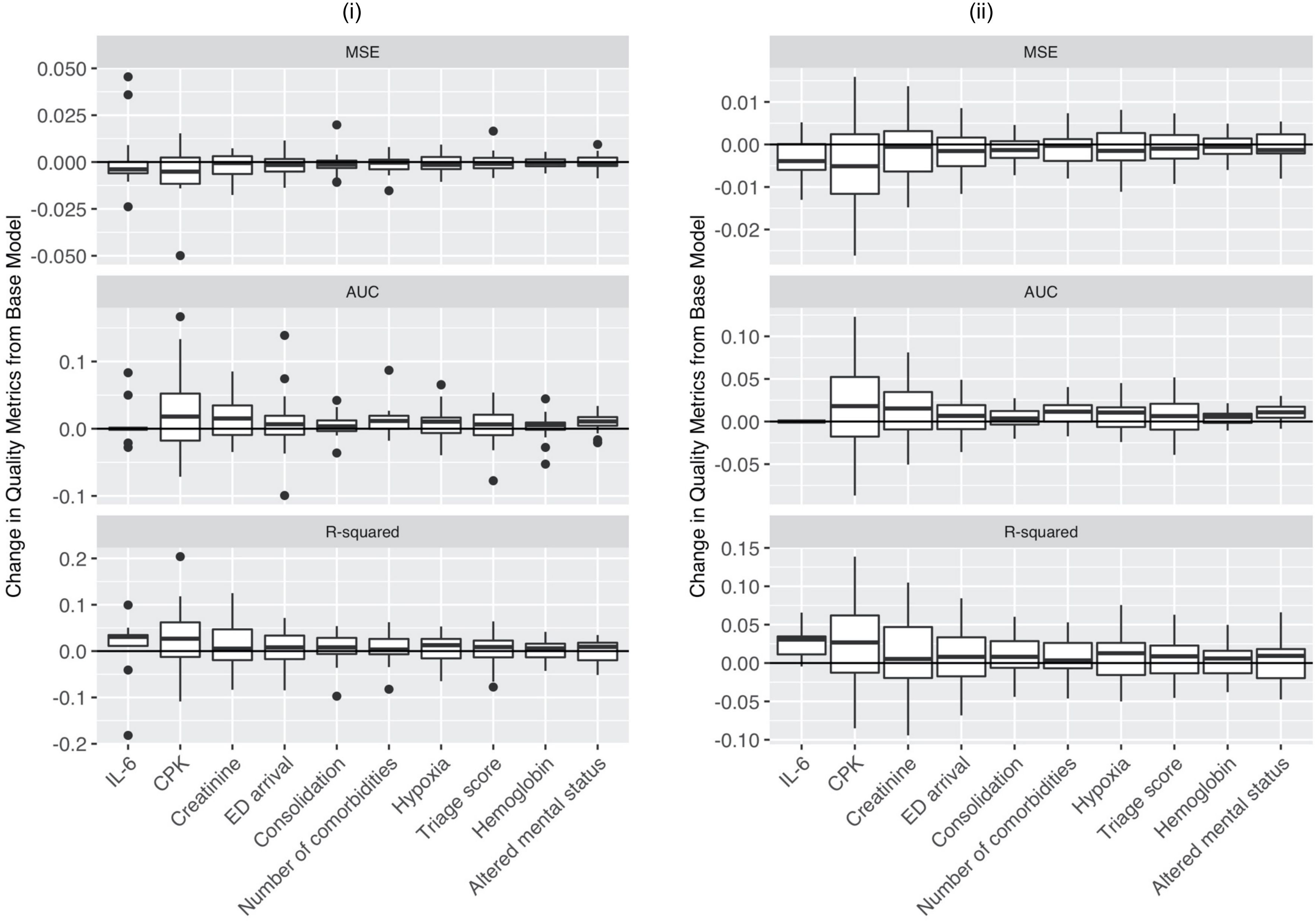

(iii)

| Variable | N | Change from Base Model<br>Mortality ~ Age + Respiratory Rate + Pulse Oximetry + Heart Rate + Hospital<br>MSE | AUC$^{(w)}$ | R-squared |
|---|---|---|---|---|
| Highest interleukin 6 (IL-6) | 130 | -0.003 | 0.001 | 0.016 |
| Creatine Phosphokinase (CPK) | 438 | -0.003 | 0.031 | 0.016 |
| **Highest creatinine** | **1687** | **-0.002** | **0.016** | **0.015** |
| ED arrival | 1610 | -0.002 | 0.007 | 0.012 |
| Consolidation on chest x-ray | 1634 | -0.001 | 0.006 | 0.007 |
| Number of comorbidities | 1716 | -0.001 | 0.013 | 0.007 |
| Hypoxia | 1716 | -0.001 | 0.011 | 0.006 |
| Triage score | 1458 | -0.001 | 0.007 | 0.005 |
| Highest hemoglobin (Hgb) | 1698 | -0.001 | 0.002 | 0.004 |
| Altered mental status | 1716 | -0.001 | 0.009 | 0.004 |

**eFigure 3c: Change in quality metrics for top 10 variables with most improved MSE when added to base model of in-hospital mortality from COVID-19 on patient's age, respiratory rate on presentation, pulse oximetry on presentation, heart rate on presentation, and hospital COVID-19 mortality rate.** (i) Histograms of change in MSE, AUC$^{(w)}$ and R-squared for all 20 hospitals in the derivation set. (ii) Histograms of change in MSE, AUC$^{(w)}$ and R-squared for all 20 hospitals in the derivation set with outliers removed. (iii) Change in quality metrics for all derivation hospitals combined. Bolded variables were chosen for inclusion in the next step of forward selection.

Based on these results, we included the patient's initial *creatinine* level in the model. The IL-6 and CPK lab values were available for very few patients. Creatinine was the factor with the most improvement in MSE and AUC$^{(w)}$ overall (iii) after these variables and it showed reasonably consistent improvement across hospitals (i and ii).

**FORWARD SELECTION STEP 4**

**Base Model: Mortality ~ Age + Respiratory Rate + Pulse Oximetry + Heart Rate + Creatinine + Hospital**

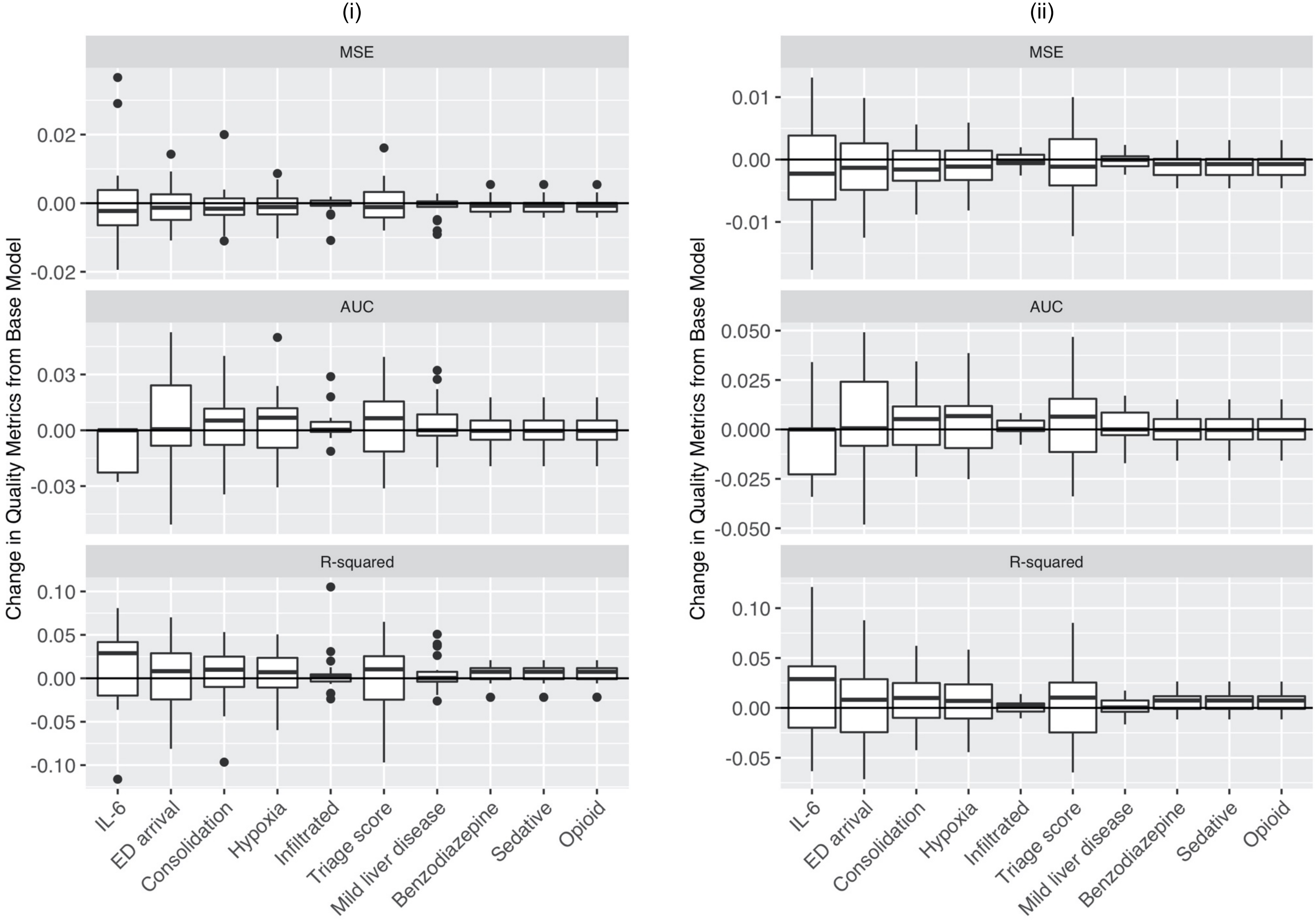

(iii)

| Variable | N | Change from Base Model<br>Mortality ~ Age + Respiratory Rate + Pulse Oximetry + Heart Rate + Creatinine + Hospital<br>MSE | AUC$^{(w)}$ | R-squared |
|---|---|---|---|---|
| Highest interleukin 6 (IL-6) | 129 | -0.003 | -0.015 | 0.017 |
| ED arrival | 1582 | -0.001 | 0.004 | 0.008 |
| Consolidation on chest x-ray | 1609 | -0.001 | 0.005 | 0.007 |
| Hypoxia | 1687 | -0.001 | 0.005 | 0.006 |
| New or worsening infiltrates on chest-xray | 1609 | -0.001 | 0.003 | 0.004 |
| Triage score | 1434 | -0.001 | 0.004 | 0.004 |
| Mild liver disease | 1687 | -0.001 | 0.002 | 0.004 |
| Previously treated by a benzodiazepine | 1687 | -0.001 | -0.001 | 0.004 |
| Previoiusly treated by a sedative | 1687 | -0.001 | -0.001 | 0.004 |
| Previoiusly treated by an opioid | 1687 | -0.001 | -0.001 | 0.004 |

**eFigure 3d: Change in quality metrics for top 10 variables with most improved MSE when added to base model of in-hospital mortality from COVID-19 on patient's age, respiratory rate on presentation, pulse oximetry on presentation, heart rate on presentation, creatinine on presentation, and hospital COVID-19 mortality rate.** (i) Histograms of change in MSE, AUC$^{(w)}$ and R-squared for all 20 hospitals in the derivation set. (ii) Histograms of change in MSE, AUC$^{(w)}$ and R-squared for all 20 hospitals in the derivation set with outliers removed. (iii) Change in quality metrics for all derivation hospitals combined.

Based on these results, we did not include any further variables in the model. Th IL-6 lab values again were available for very few patients. After that, none of the other variables improved MSE, AUC$^{(w)}$, or R-squared overall enough to warrant inclusion (iii).

**BACKWARD SELECTION STEP 1**

**Base Model: Mortality ~ Age + Respiratory Rate + Pulse Oximetry + Heart Rate + Creatinine + Hospital**

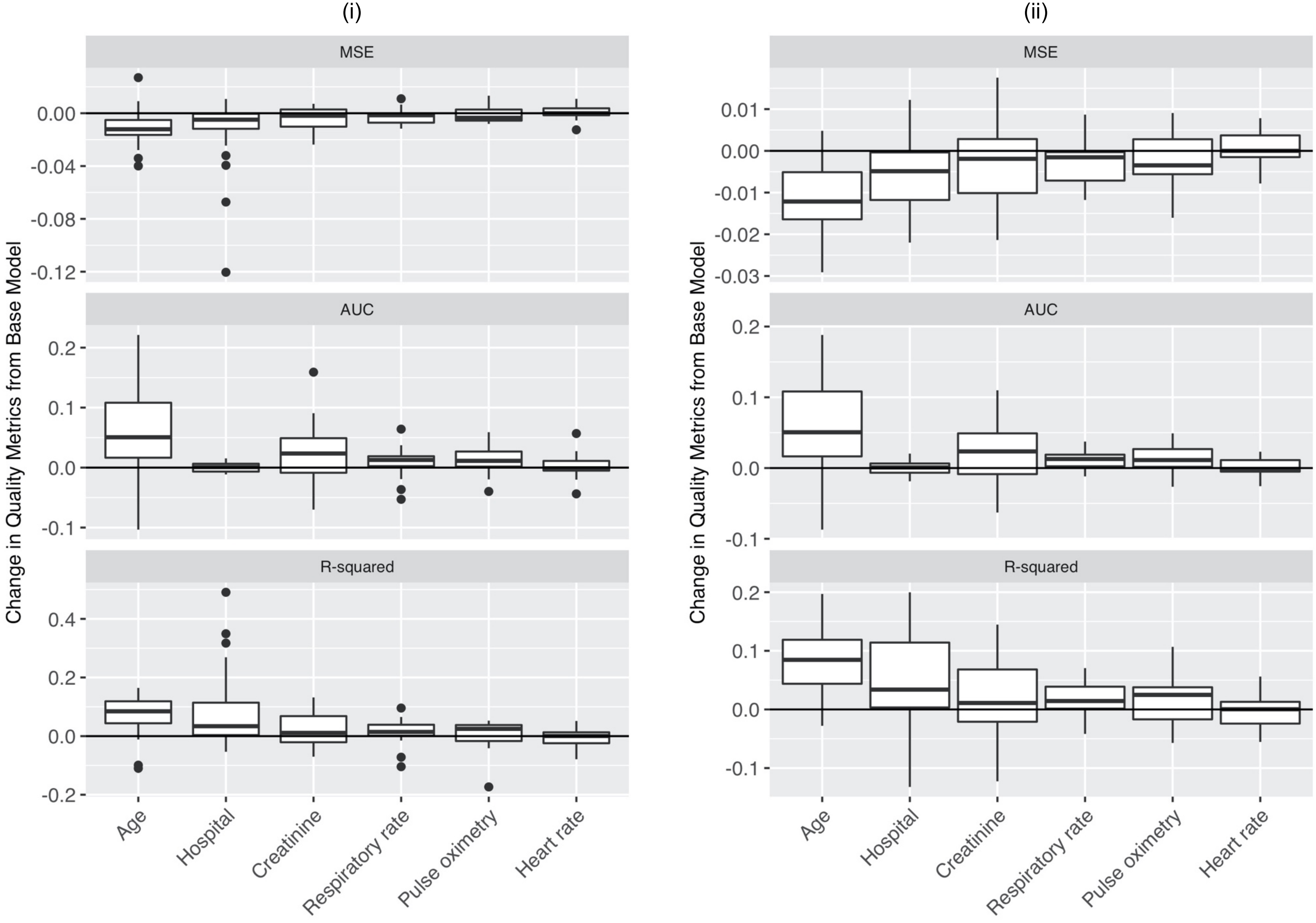

**eFigure 3e: Change in quality metrics for backward selection when removed from base model of in-hospital mortality from COVID-19 on patient's age, respiratory rate on presentation, pulse oximetry on presentation, heart rate on presentation, creatinine on presentation, and hospital COVID-19 mortality rate.** (i) Histograms of change in MSE, $AUC^{(w)}$ and R-squared for all 20 hospitals in the derivation set. (ii) Histograms of change in MSE, $AUC^{(w)}$ and R-squared for all 20 hospitals in the derivation set with outliers removed.

Based on these results, we decided to remove heart rate on presentation from the model because removing it from the model actually consistently improved MSE and R-squared across individual hospitals (i and ii). All other variables were kept in the model, because removing them from the model hurt the quality metrics across individual hospitals.

**FORWARD SELECTION STEP 5**
**Base Model: Mortality ~ Age + Respiratory Rate + Pulse Oximetry + Creatinine + Hospital**

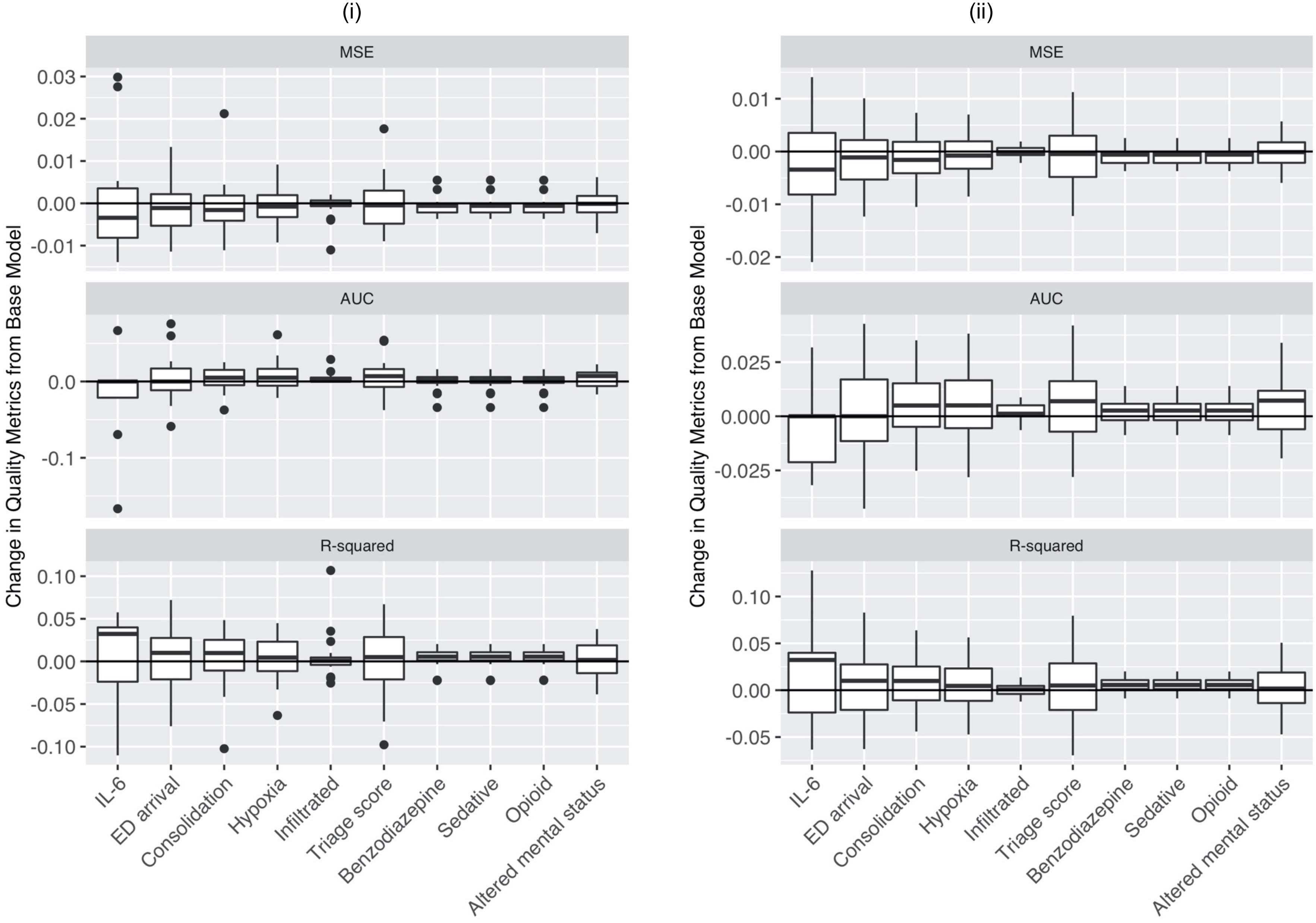

(iii)

| Variable | N | Change from Base Model Mortality ~ Age + Respiratory Rate + Pulse Oximetry + Creatinine + Hospital | | |
|---|---|---|---|---|
| | | MSE | AUC(w) | R-squared |
| Highest interleukin 6 (IL-6) | 130 | -0.004 | -0.017 | 0.021 |
| ED arrival | 1585 | -0.001 | 0.004 | 0.009 |
| Consolidation on chest x-ray | 1611 | -0.001 | 0.004 | 0.006 |
| Hypoxia | 1690 | -0.001 | 0.008 | 0.005 |
| New or worsening infiltrates on chest-xray | 1611 | -0.001 | 0.004 | 0.004 |
| Triage score | 1437 | -0.001 | 0.007 | 0.004 |
| Previously treated by a benzodiazepine | 1690 | -0.001 | <0.001 | 0.003 |
| Previoiusly treated by a sedative | 1690 | -0.001 | <0.001 | 0.003 |
| Previoiusly treated by an opioid | 1690 | -0.001 | <0.001 | 0.003 |
| Altered mental status | 1690 | <0.001 | 0.004 | 0.003 |

**eFigure 3f: Change in quality metrics for top 10 variables with most improved MSE when added to base model of in-hospital mortality from COVID-19 on patient's age, respiratory rate on presentation, pulse oximetry on presentation, creatinine on presentation, and hospital COVID-19 mortality rate.** (i) Histograms of change in MSE, AUC(w) and R-squared for all 20 hospitals in the derivation set. (ii) Histograms of change in MSE, AUC(w) and R-squared for all 20 hospitals in the derivation set with outliers removed. (iii) Change in quality metrics for all derivation hospitals combined.

Based on these results, we did not include any further variables in the model. The IL-6 lab values again were available for very few patients. After that, none of the other variables improved MSE, AUC(w), or R-squared overall enough to warrant inclusion (iii).

### eAppendix B: Model discrimination within subgroups

In order to understand how the model performed for patients with different characteristics (fairness), we calculated the model AUC with the full data (derivation and validation set combined), but subsetted to different subgroups of race, gender, and age. Because the median age of patients who died in the data was 74 years, we use this as a cutoff for the age subgroup analysis. We note that the model shows similar discrimination for Black and white patients, as well as for female and male patients (Appendix Table 4). The model shows less discrimination for patients 75 years or older, but still improves over random guessing by almost 20% (Appendix Table 4).

(A)

| | Race | |
|---|---|---|
| | **Black** | **White** |
| AUC by group | 0.80 [n=966] | 0.78 [n=909] |
| Overall AUC | 0.79 [N = 1,875] | |

(B)

| | Gender | |
|---|---|---|
| | **Female** | **Male** |
| AUC by group | 0.81 [n=1,000] | 0.79 [n=1,088] |
| Overall AUC | 0.80 [N = 2,088] | |

(C)

| | Age | |
|---|---|---|
| | **< 75** | **75 or older** |
| AUC by group | 0.80 [n=1,503] | 0.68 [n=585] |
| Overall AUC | 0.80 [N = 2,088] | |

AUC values are calculated with predictions assuming a hospital mortality rate of .2 for all patients, using the full complete cases dataset (derivation and validation sets combined with complete cases for the variables included in the risk model).
The subgroup analysis for race excludes individuals who were not identified as either Black or white in the dataset, and therefore the overall AUC is calculated with a subset of this data.

### eTable 4: AUC by subgroup. (A) Race. (B) Gender. (C) Age.
